# Systemic SARS-CoV-2-specific antibody responses to infection and to COVID-19 and BCG vaccination

**DOI:** 10.1101/2024.01.24.24301644

**Authors:** Juana Claus, Thijs ten Doesschate, Esther Taks, Priya Debisarun, Gaby Smits, Rob van Binnendijk, Fiona van der Klis, Lilly M. Verhagen, Marien I. de Jonge, Marc J.M. Bonten, Mihai G. Netea, Janneke H. H. M. van de Wijgert

**Affiliations:** Julius Center for Health Sciences and Primary Care, University Medical Center Utrecht, Utrecht University, Utrecht, Netherlands; Department of Internal Medicine, Jeroen Bosch Ziekenhuis, ‘s Hertogenbosch, Netherlands; Department of Medicine and Radboud Center for Infectious Diseases, Radboud University Medical Center, Nijmegen, the Netherlands; National Institute of Public Health and the Environment, Bilthoven, Netherlands; Department of Paediatric Infectious Diseases and Immunology, Amalia Children’s Hospital, Radboud University Medical Center, Nijmegen, Netherlands; Department of Laboratory Medicine, Laboratory of Medical Immunology, Radboud Center for Infectious Diseases, Radboud University Medical Center, Nijmegen, Netherlands; Department for Genomics & Immunoregulation, Life and Medical Sciences Institute, University of Bonn, Germany

**Keywords:** SARS-CoV-2, COVID-19, serology, infection, vaccination, BCG

## Abstract

SARS-CoV-2 infections elicit antibodies against the viral spike (S) and nucleocapsid (N) proteins; COVID-19 vaccines against the S-protein only. The BCG-Corona trial, initiated in March 2020 in SARS-CoV-2-naïve Dutch healthcare workers, captured several epidemic peaks and the introduction of COVID-19 vaccines during the one-year follow-up. We assessed determinants of systemic anti-S1 and anti-N immunoglobulin type G (IgG) responses using trial data. Participants were randomized to BCG or placebo vaccination, reported daily symptoms, SARS-CoV-2 test results, and COVID-19 vaccinations, and donated blood for SARS-CoV-2 serology at two time points. In the 970 participants, anti-S1 geometric mean antibody concentrations (GMCs) were much higher than anti-N GMCs. Anti-S1 GMCs significantly increased with increasing number of immune events (SARS-CoV-2 infection or COVID-19 vaccination): 104.7 international units (IU)/ml, 955.0 IU/ml, and 2290.9 IU/ml for one, two, and three immune events, respectively (p<0.001). In adjusted multivariable linear regression models, anti-S1 and anti-N log_10_ concentrations were significantly associated with infection severity, and anti-S1 log_10_ concentration with COVID-19 vaccine type/dose. In univariable models, anti-N log_10_ concentration was also significantly associated with acute infection duration, and severity and duration of individual symptoms. Antibody concentrations were not associated with Long COVID or long-term loss of smell/taste.

## Introduction

The systemic humoral immune response to SARS-CoV-2 is characterized by antigen-specific antibodies, which play a crucial role in host defence and protection against re-infection. Following an infection, systemic immunoglobulins type G (IgG) specific to SARS-CoV-2, including antibodies against the viral spike (S) and nucleocapsid (N) proteins, are detected in the majority of patients within two weeks, with peak concentrations at around three weeks after symptom onset [1–3]. While natural infection typically gives rise to both anti-S and anti-N antibodies, the COVID-19 vaccines that were authorized in the European Union within the first two years of the epidemic (mRNA [4, 5] and viral vector vaccines[6, 7]) were engineered to elicit an anti-S immune response only [3]. The presence of anti-S antibodies is a more sensitive and specific marker for the presence of a SARS-CoV-2 infection than the presence of anti-N antibodies[8], with a reported sensitivity of 91.3-97.1% (depending on infection severity) and specificity of 98.1% [9]. Anti-N sensitivities are estimated to be 85% for mild infections and 67% for asymptomatic infections, and specificity 97% (96%-98%) [10].

The S-protein, containing the S1 subunit with the receptor binding domain, is abundant and readily accessible on the viral surface. Conversely, the N-protein resides within the core of the viral structure and is less exposed to the host immune system [3]. Furthermore, the N-protein is highly conserved among the different coronaviruses, including some common cold-causing coronaviruses that circulate widely, and existing immunity to anti-N is therefore present in most populations [11, 12]. However, anti-N is the only biomarker currently available to detect recent natural infections in a vaccinated population [10].

Studies have suggested that antibody concentrations over time likely vary by numbers, types, and combinations of immune events (SARS-CoV-2 infections or COVID-19 vaccinations), disease severity, and other virus, host, and vaccine characteristics [13–16]. However, asymptomatic and very mild cases in relatively healthy hosts were underrepresented in studies thus far, with most research focused on the broad WHO-defined mild, moderate, and severe infections [17]. We therefore used data from our previously published BCG-Corona trial in healthy healthcare workers [18, 19] to assess systemic anti-S1 IgG and anti-N IgG responses at the end of the first year of the SARS-CoV-2 epidemic in the Netherlands. Previous BCG-Corona trial analyses found that baseline BCG vaccination did not reduce SARS-CoV-2 infection incidence, duration, or severity, nor SARS-CoV-2 antibody responses, in the first year after vaccination [19]. In this paper, our aim was to assess whether numbers, types, and combinations of immune events, SARS-CoV-2 infection severity and duration, and participant characteristics (including having received BCG or placebo vaccination at baseline) were associated with systemic SARS-CoV-2-specific antibody concentrations after one year of follow-up. The strengths of these analyses include coverage of the first year of the epidemic when participants were still immunologically naïve for SARS-CoV-2 and the completeness of the dataset. BCG-Corona study participants reported symptoms daily, and positive SARS-CoV-2 test results, healthcare seeking behaviour, and vaccinations weekly, and donated blood samples for serology twice in one year. This comprehensive data enabled us to characterize infections, including asymptomatic and very mild infections, in great detail.

## Methods

### Study design and participants

The BCG-Corona trial was a multi-centre, double-blind, placebo-controlled randomized trial. The study protocol was approved by the institutional review board of the University Medical Center Utrecht, the Netherlands, and, registered at clinicaltrials.gov (identifier: NCT04328441).[20] The trial protocol and primary results of the trial have been published [18, 19]. Healthcare workers from nine Dutch hospitals who were expected to be in direct contact with COVID-19 patients were vaccinated with BCG or placebo vaccine (1:1) during the first SARS-CoV-2 epidemic wave in March/April 2020 and followed up for about one year. The participating study sites were categorized into core hospitals (N=3) that implemented on-site venepuncture sampling, and other hospitals (N=6) that implemented at-home fingerprick sampling (full eligibility criteria and sampling procedures in Supplementary Material, available on the Cambridge Core website).

### Data collection and seroconversion periods

Participants were invited to donate a blood sample at two time points during follow-up. This divided the follow-up time into two potential seroconversion periods. The first seroconversion period started on the date of randomisation until the date of the first sampling round, about three months (M3) following BCG/placebo vaccination for the core hospitals and about six months (M6) for the other hospitals. The second seroconversion period was between the first sampling round and the second sampling round, approximately 12 months (M12) after BCG/placebo vaccination for all hospitals. Total follow-up time for each participant was the number of days between randomisation and the date of M12 sampling. Baseline characteristics were entered into an online questionnaire by study staff at the time of randomisation (Research Online, Julius Center, UMC Utrecht, the Netherlands). Participants were instructed to report daily symptoms including their severity using a smartphone diary application (Research Follow App, Your Research BV, Huizen, Netherlands) as well as the dates and results of any SARS-CoV-2 test, any other healthcare seeking behaviour, and any vaccination (Supplementary Methods).

### COVID-19 vaccinations

COVID-19 vaccine roll-out in the Netherlands began on 6 January 2021, and the diary app was updated with weekly questions about the date(s) and types of COVID-19 vaccinations at this time. The COVID-19 vaccines available during the study period were the mRNA vaccines Spikevax (Moderna Biotech, Cambridge, MA, USA) and Comirnaty (Pfizer/BioNTech, New York, NY, USA), and the vector vaccines Vaxzevria (AstraZeneca AB, Sodertalje, Sweden) and Jcovden (Janssen Vaccines, Leiden, Netherlands). By the time the M12 blood sampling was completed in June 2021, healthcare workers could have received zero, one or two doses of only one vaccine type.

### Antibody measurements

All systemic antibody testing was conducted by the Center for Immunology of Infections and Vaccines at the National Institute for Public Health and the Environment in Bilthoven, the Netherlands. In the first seroconversion period, prior to the start of the COVID-19 vaccination campaign, only anti-S1 IgG antibodies were assessed. At M12, both anti-S1 and anti-N IgG antibodies were assessed to enable differentiation between natural infections and vaccine-induced antibodies. Anti-S1 and anti-N concentrations were measured using an in-house magnetic immunoassay on a Luminex platform, with reported sensitivities of 91.3-97.1% and 67-85% (depending on infection severity) for anti-S1 and anti-N respectively, and specificities of 98.1% and 97%, respectively [9, 10]. Anti-S1 and anti-N concentrations were measured simultaneously if applicable. The concentrations were subsequently calibrated against the international standard for human anti-SARS-CoV-2 immunoglobulin (20/136 NIBSC standard [21]) and expressed as international units per mL (IU/mL)[10, 22]. The threshold for seropositivity was set to 10.1 IU/mL for anti-S1 and 14.3 IU/mL for anti-N. Samples that were below these thresholds were considered to have concentrations of zero. The anti-S1 and anti-N concentrations displayed gamma distributions and were log_10_ transformed after adding a pseudocount of +1 to each value to eliminate zero values (Figure S1). We focused the analyses on the M12 antibody concentrations, because most immune events took place between the first (M3/M6) and second (M12) sampling rounds.

### Definitions of antibody concentration determinants

A single immune event was defined as one SARS-CoV-2 infection episode or one dose of a COVID-19 vaccine. An infection episode was defined as a positive test (PCR or rapid antigen test) reported by the participant and/or evidence of seroconversion (for anti-S1 in the first seroconversion period and anti-S1 plus anti-N in the second seroconversion period). We also categorized participants by the vaccination types and doses that they received, and immunity type: natural immunity, vaccine immunity (after one or two doses), or hybrid immunity (a combination of natural infection and one or two vaccine doses).

To determine infection severity, we matched each infection episode with corresponding self-reported symptoms and healthcare seeking behaviour information (Supplementary Methods). We subsequently categorized each infection episode in accordance with World Health Organisation (WHO) severity categories of asymptomatic, mild, moderate and severe [17]. However, only three BCG-Corona participants were hospitalized (WHO moderate category), and all other symptomatic episodes qualified as WHO mild. We therefore further subdivided the WHO mild category into very mild and mild subcategories (Supplementary Methods): mild episodes were accompanied by more severe respiratory symptoms and/or fever, and lasted longer, compared to very mild episodes. The duration of the acute phase of the infection episode was defined as the number of consecutive days during which the participant reported symptoms, not including standalone loss of smell/taste. Long-term loss of smell/taste and Long COVID (lingering symptoms other than standalone loss of smell/taste) were defined as continuing to report the respective symptoms for at least 60 days after the end of the acute infection episode. Duration of episodes that were ongoing at the end of follow-up were coded as unknown. Chronic symptoms that were reported regularly throughout follow-up were ignored.

### Statistical analyses

Statistical analyses were performed in R version 4.2.2 (PBC, Boston, MA, USA). Participants were excluded from the analysis population if they did not donate a M12 sample, their infection status was unsure, or their sample was taken in a seroconversion window after infection or vaccination. Infection status was considered unsure if a participant never reported a positive test but had completed less than 80% of diary entries, or had anti-N but no anti-S1 antibodies at M12. The seroconversion window was defined as 14 days, but sensitivity analyses were conducted using 7 and 0 days. Participants who were in the seroconversion window for the second dose of a COVID-19 vaccine were included as having received only one dose.

Characteristics between groups were compared using Chi-squared test for categorical variables and Wilcoxon rank sum test for continuous variables. The antilog 10^x^ of mean concentrations was taken to obtain geometric mean concentrations (GMCs). Mean log10-transformed antibody concentrations between groups were compared using Kruskal-Wallis tests. Associations between covariates (with categorical variables analysed as indicator variables) and log10-transformed anti-S1 and anti-N concentrations at M12 were assessed using linear regression models. BCG versus placebo vaccination at baseline, age, and sex were forced into each multivariable model. Other covariates were considered for inclusion in multivariable models based on statistical significance in univariable linear regression, using p<0.05 as the cut-off value. To avoid multicollinearity, we included combination variables for COVID-19 vaccination (number of doses received plus vaccine product if at least one dose received) and overall infection severity (whether an infection took place and if yes, its overall severity based on both symptom severity and duration), but not number of immune events, overall acute infection episode duration, or severity/duration of individual symptoms.

## Results

### Participant characteristics

The analysis population consisted of 970 of the 1511 randomized participants (Figure 1). Baseline characteristics were comparable between the two populations, and between the BCG and placebo groups within each population (Table 1A) [19]. The majority of the participants (74.7%) were female, with a mean age of 42.5 years (range 18-67), and had high (60%) to medium risk (24.7%) of work-related SARS-CoV-2 exposure (Supplementary Methods). The prevalence of chronic comorbidities other than hay fever was low. As expected, immune event-related characteristics during follow-up differed between the analysis and randomized populations (Table 1B) because application of the 14-day seroconversion window resulted in the removal of some immune events from the analysis population (Figure S2; Table S1).

**Figure 1:**
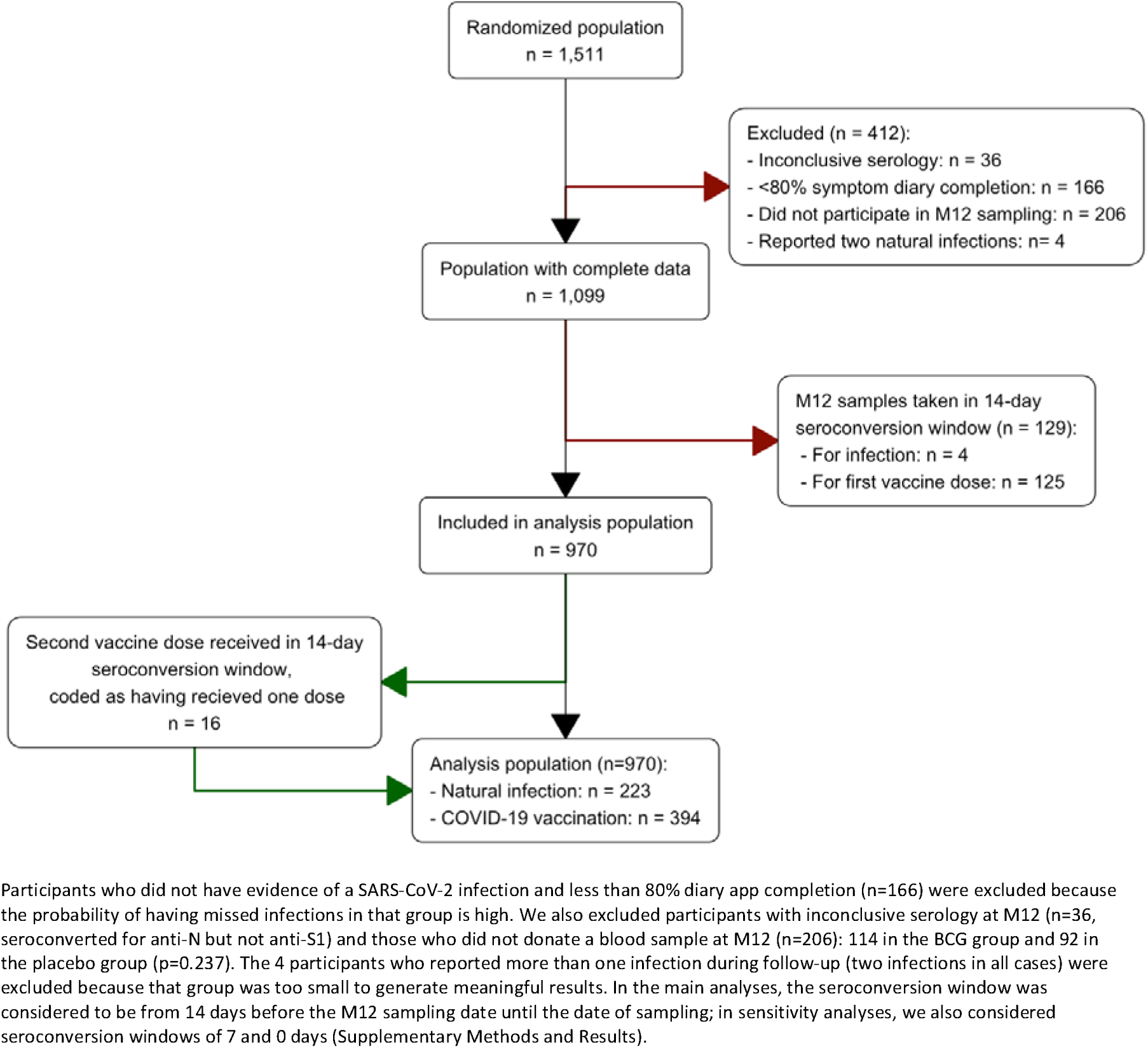
Population and study flow.

**Table 1:**
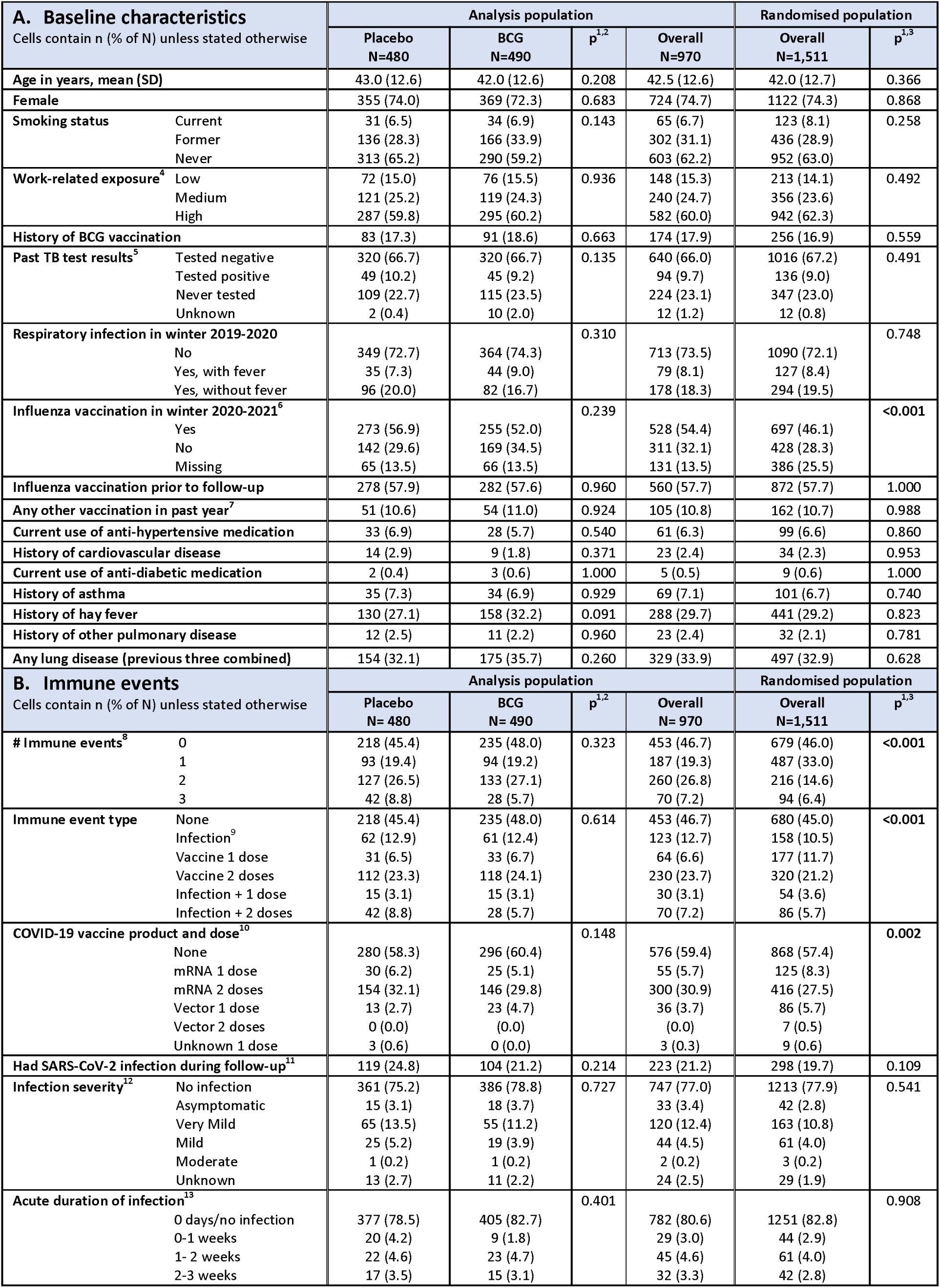
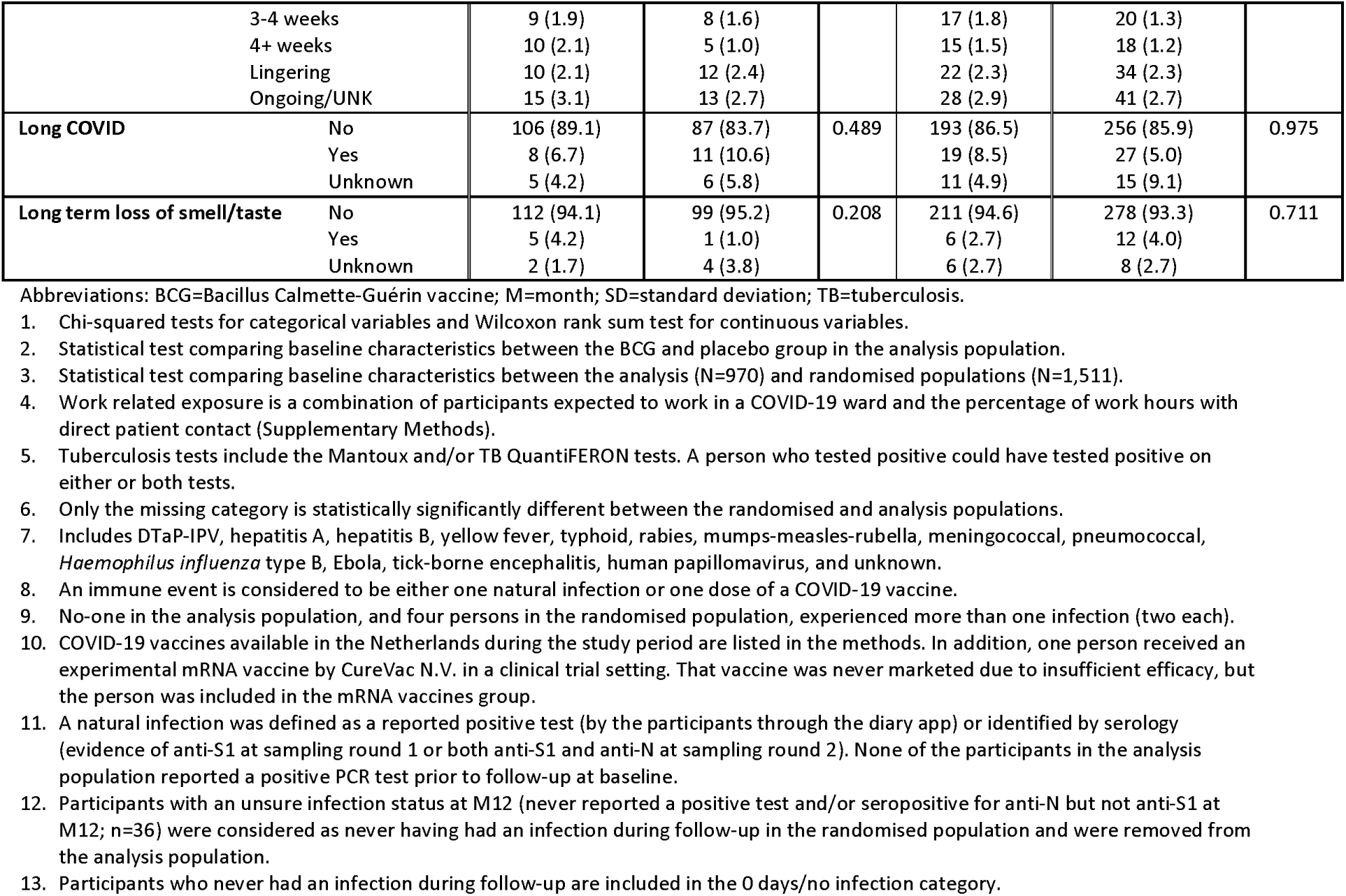
Participant baseline characteristics and immune events during follow-up.

| **A. Baseline characteristics**<br>Cells contain n (% of N) unless stated otherwise |  | Analysis population |  |  |  | Randomised population |  |
| --- | --- | --- | --- | --- | --- | --- | --- |
|  |  | Placebo<br>N=480 | BCG<br>N=490 | p<sup>1,2</sup> | Overall<br>N=970 | Overall<br>N=1,511 | p<sup>1,3</sup> |
| Age in years, mean (SD) |  | 43.0 (12.6) | 42.0 (12.6) | 0.208 | 42.5 (12.6) | 42.0 (12.7) | 0.366 |
| Female |  | 355 (74.0) | 369 (72.3) | 0.683 | 724 (74.7) | 1122 (74.3) | 0.868 |
| Smoking status | Current | 31 (6.5) | 34 (6.9) | 0.143 | 65 (6.7) | 123 (8.1) | 0.258 |
|  | Former | 136 (28.3) | 166 (33.9) |  | 302 (31.1) | 436 (28.9) |  |
|  | Never | 313 (65.2) | 290 (59.2) |  | 603 (62.2) | 952 (63.0) |  |
| Work-related exposure<sup>4</sup> | Low | 72 (15.0) | 76 (15.5) | 0.936 | 148 (15.3) | 213 (14.1) | 0.492 |
|  | Medium | 121 (25.2) | 119 (24.3) |  | 240 (24.7) | 356 (23.6) |  |
|  | High | 287 (59.8) | 295 (60.2) |  | 582 (60.0) | 942 (62.3) |  |
| History of BCG vaccination |  | 83 (17.3) | 91 (18.6) | 0.663 | 174 (17.9) | 256 (16.9) | 0.559 |
| Past TB test results<sup>5</sup> | Tested negative | 320 (66.7) | 320 (66.7) | 0.135 | 640 (66.0) | 1016 (67.2) | 0.491 |
|  | Tested positive | 49 (10.2) | 45 (9.2) |  | 94 (9.7) | 136 (9.0) |  |
|  | Never tested | 109 (22.7) | 115 (23.5) |  | 224 (23.1) | 347 (23.0) |  |
|  | Unknown | 2 (0.4) | 10 (2.0) |  | 12 (1.2) | 12 (0.8) |  |
| Respiratory infection in winter 2019-2020 |  |  |  | 0.310 |  |  | 0.748 |
|  | No | 349 (72.7) | 364 (74.3) |  | 713 (73.5) | 1090 (72.1) |  |
|  | Yes, with fever | 35 (7.3) | 44 (9.0) |  | 79 (8.1) | 127 (8.4) |  |
|  | Yes, without fever | 96 (20.0) | 82 (16.7) |  | 178 (18.3) | 294 (19.5) |  |
| Influenza vaccination in winter 2020-2021<sup>6</sup> |  |  |  | 0.239 |  |  | <0.001 |
|  | Yes | 273 (56.9) | 255 (52.0) |  | 528 (54.4) | 697 (46.1) |  |
|  | No | 142 (29.6) | 169 (34.5) |  | 311 (32.1) | 428 (28.3) |  |
|  | Missing | 65 (13.5) | 66 (13.5) |  | 131 (13.5) | 386 (25.5) |  |
| Influenza vaccination prior to follow-up |  | 278 (57.9) | 282 (57.6) | 0.960 | 560 (57.7) | 872 (57.7) | 1.000 |
| Any other vaccination in past year<sup>7</sup> |  | 51 (10.6) | 54 (11.0) | 0.924 | 105 (10.8) | 162 (10.7) | 0.988 |
| Current use of anti-hypertensive medication |  | 33 (6.9) | 28 (5.7) | 0.540 | 61 (6.3) | 99 (6.6) | 0.860 |
| History of cardiovascular disease |  | 14 (2.9) | 9 (1.8) | 0.371 | 23 (2.4) | 34 (2.3) | 0.953 |
| Current use of anti-diabetic medication |  | 2 (0.4) | 3 (0.6) | 1.000 | 5 (0.5) | 9 (0.6) | 1.000 |
| History of asthma |  | 35 (7.3) | 34 (6.9) | 0.929 | 69 (7.1) | 101 (6.7) | 0.740 |
| History of hay fever |  | 130 (27.1) | 158 (32.2) | 0.091 | 288 (29.7) | 441 (29.2) | 0.823 |
| History of other pulmonary disease |  | 12 (2.5) | 11 (2.2) | 0.960 | 23 (2.4) | 32 (2.1) | 0.781 |
| Any lung disease (previous three combined) |  | 154 (32.1) | 175 (35.7) | 0.260 | 329 (33.9) | 497 (32.9) | 0.628 |
| **B. Immune events**<br>Cells contain n (% of N) unless stated otherwise |  | Analysis population |  |  |  | Randomised population |  |
|  |  | Placebo<br>N= 480 | BCG<br>N= 490 | p<sup>1,2</sup> | Overall<br>N= 970 | Overall<br>N=1,511 | p<sup>1,3</sup> |
| # Immune events<sup>8</sup> | 0 | 218 (45.4) | 235 (48.0) | 0.323 | 453 (46.7) | 679 (46.0) | <0.001 |
|  | 1 | 93 (19.4) | 94 (19.2) |  | 187 (19.3) | 487 (33.0) |  |
|  | 2 | 127 (26.5) | 133 (27.1) |  | 260 (26.8) | 216 (14.6) |  |
|  | 3 | 42 (8.8) | 28 (5.7) |  | 70 (7.2) | 94 (6.4) |  |
| Immune event type | None | 218 (45.4) | 235 (48.0) | 0.614 | 453 (46.7) | 680 (45.0) | <0.001 |
|  | Infection<sup>9</sup> | 62 (12.9) | 61 (12.4) |  | 123 (12.7) | 158 (10.5) |  |
|  | Vaccine 1 dose | 31 (6.5) | 33 (6.7) |  | 64 (6.6) | 177 (11.7) |  |
|  | Vaccine 2 doses | 112 (23.3) | 118 (24.1) |  | 230 (23.7) | 320 (21.2) |  |
|  | Infection + 1 dose | 15 (3.1) | 15 (3.1) |  | 30 (3.1) | 54 (3.6) |  |
|  | Infection + 2 doses | 42 (8.8) | 28 (5.7) |  | 70 (7.2) | 86 (5.7) |  |
| COVID-19 vaccine product and dose<sup>10</sup> |  |  |  | 0.148 |  |  | 0.002 |
|  | None | 280 (58.3) | 296 (60.4) |  | 576 (59.4) | 868 (57.4) |  |
|  | mRNA 1 dose | 30 (6.2) | 25 (5.1) |  | 55 (5.7) | 125 (8.3) |  |
|  | mRNA 2 doses | 154 (32.1) | 146 (29.8) |  | 300 (30.9) | 416 (27.5) |  |
|  | Vector 1 dose | 13 (2.7) | 23 (4.7) |  | 36 (3.7) | 86 (5.7) |  |
|  | Vector 2 doses | 0 (0.0) | 0 (0.0) |  | 0 (0.0) | 7 (0.5) |  |
|  | Unknown 1 dose | 3 (0.6) | 0 (0.0) |  | 3 (0.3) | 9 (0.6) |  |
| Had SARS-CoV-2 infection during follow-up<sup>11</sup> |  | 119 (24.8) | 104 (21.2) | 0.214 | 223 (21.2) | 298 (19.7) | 0.109 |
| Infection severity<sup>12</sup> | No infection | 361 (75.2) | 386 (78.8) | 0.727 | 747 (77.0) | 1213 (77.9) | 0.541 |
|  | Asymptomatic | 15 (3.1) | 18 (3.7) |  | 33 (3.4) | 42 (2.8) |  |
|  | Very Mild | 65 (13.5) | 55 (11.2) |  | 120 (12.4) | 163 (10.8) |  |
|  | Mild | 25 (5.2) | 19 (3.9) |  | 44 (4.5) | 61 (4.0) |  |
|  | Moderate | 1 (0.2) | 1 (0.2) |  | 2 (0.2) | 3 (0.2) |  |
|  | Unknown | 13 (2.7) | 11 (2.2) |  | 24 (2.5) | 29 (1.9) |  |
| Acute duration of infection<sup>13</sup> |  |  |  | 0.401 |  |  | 0.908 |
|  | 0 days/no infection | 377 (78.5) | 405 (82.7) |  | 782 (80.6) | 1251 (82.8) |  |
|  | 0-1 weeks | 20 (4.2) | 9 (1.8) |  | 29 (3.0) | 44 (2.9) |  |
|  | 1-2 weeks | 22 (4.6) | 23 (4.7) |  | 45 (4.6) | 61 (4.0) |  |
|  | 2-3 weeks | 17 (3.5) | 15 (3.1) |  | 32 (3.3) | 42 (2.8) |  |
|  | 3-4 weeks | 9 (1.9) | 8 (1.6) |  | 17 (1.8) | 20 (1.3) |  |
|  | 4+ weeks | 10 (2.1) | 5 (1.0) |  | 15 (1.5) | 18 (1.2) |  |
|  | Lingering | 10 (2.1) | 12 (2.4) |  | 22 (2.3) | 34 (2.3) |  |
|  | Ongoing/UNK | 15 (3.1) | 13 (2.7) |  | 28 (2.9) | 41 (2.7) |  |
| Long COVID | No | 106 (89.1) | 87 (83.7) | 0.489 | 193 (86.5) | 256 (85.9) | 0.975 |
|  | Yes | 8 (6.7) | 11 (10.6) |  | 19 (8.5) | 27 (5.0) |  |
|  | Unknown | 5 (4.2) | 6 (5.8) |  | 11 (4.9) | 15 (9.1) |  |
| Long term loss of smell/taste | No | 112 (94.1) | 99 (95.2) | 0.208 | 211 (94.6) | 278 (93.3) | 0.711 |
|  | Yes | 5 (4.2) | 1 (1.0) |  | 6 (2.7) | 12 (4.0) |  |
|  | Unknown | 2 (1.7) | 4 (3.8) |  | 6 (2.7) | 8 (2.7) |  |

Abbreviations: BCG=Bacillus Calmette-Guérin vaccine; M=month; SD=standard deviation; TB=tuberculosis.

### M12 anti-S1 and anti-N IgG GMCs by immune events

The M12 anti-S1 and anti-N GMCs did not differ between the BCG and placebo groups (Table 2, Figures 2A-B), also not after stratification by number of immune events or immunity type (Figure S3). About half of the participants (517; 53.3%) experienced an immune event during follow up: 187 (19.3%) experienced one, 260 (26.8%) two, and 70 (7.2%) three immune events (Table 1B). About two thirds (65.8%) of the participants who experienced one immune event had an infection and 34.2% received one dose of a COVID-19 vaccine; 11.5% of the participants who experienced two immune events had an infection plus received one dose of a COVID-19 vaccine, while 88.5% received two doses of a COVID-19 vaccine. All participants with three immune events had an infection plus received two doses of a COVID-19 vaccine. M12 anti-S1 GMCs were higher with increasing number of immune events: 104.7 IU/ml (standard deviation (SD)=7.2) for one, 955.0 IU/ml (SD=4.1) for two, and 2290.9 IU/ml (SD=2.6) for three immune events (p<0.001; Table 2, Figures 2C+E). The M12 anti-S1 GMC was higher after one COVID-19 vaccine dose compared to after one infection: 177.8 IU/ml (SD=7.59) and 79.4 IU/ml (SD=6.61), respectively (p=0.010). The M12 anti-N GMC increased after infection but not after vaccination(s) as expected (Table 2, Figures 2D+F), and was lower than the anti-S1 GMC after infection: 26.3 IU/ml (SD=4.5) compared to 79.4 IU/ml (SD=6.6), respectively (p<0.001). M12 anti-S1 and anti-N log_10_ concentrations by time since an immune event showed considerable inter-individual variability but not a general downward trend, suggesting the follow-up period was not long enough for antibody waning (Figure S4).

**Figure 2:**
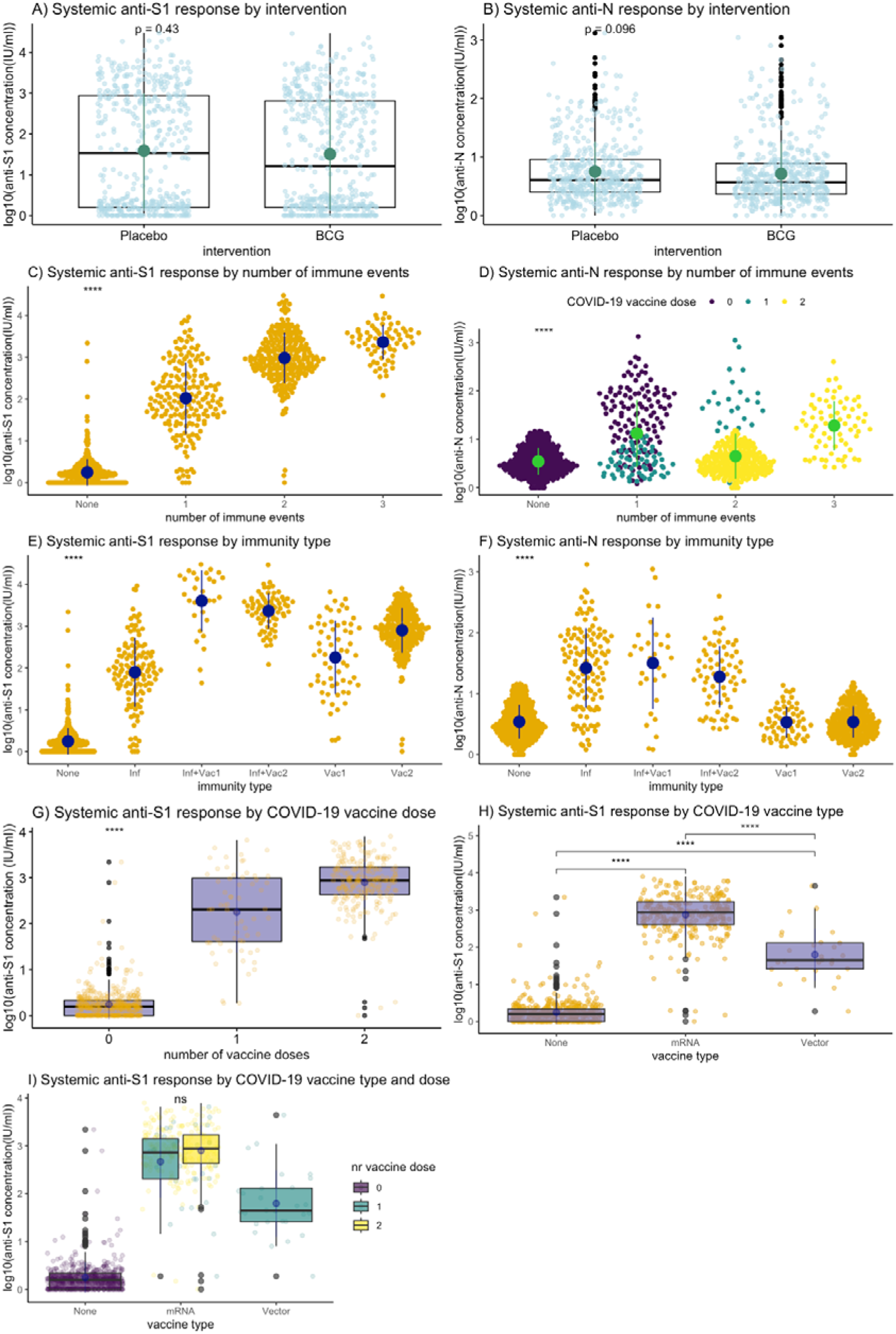
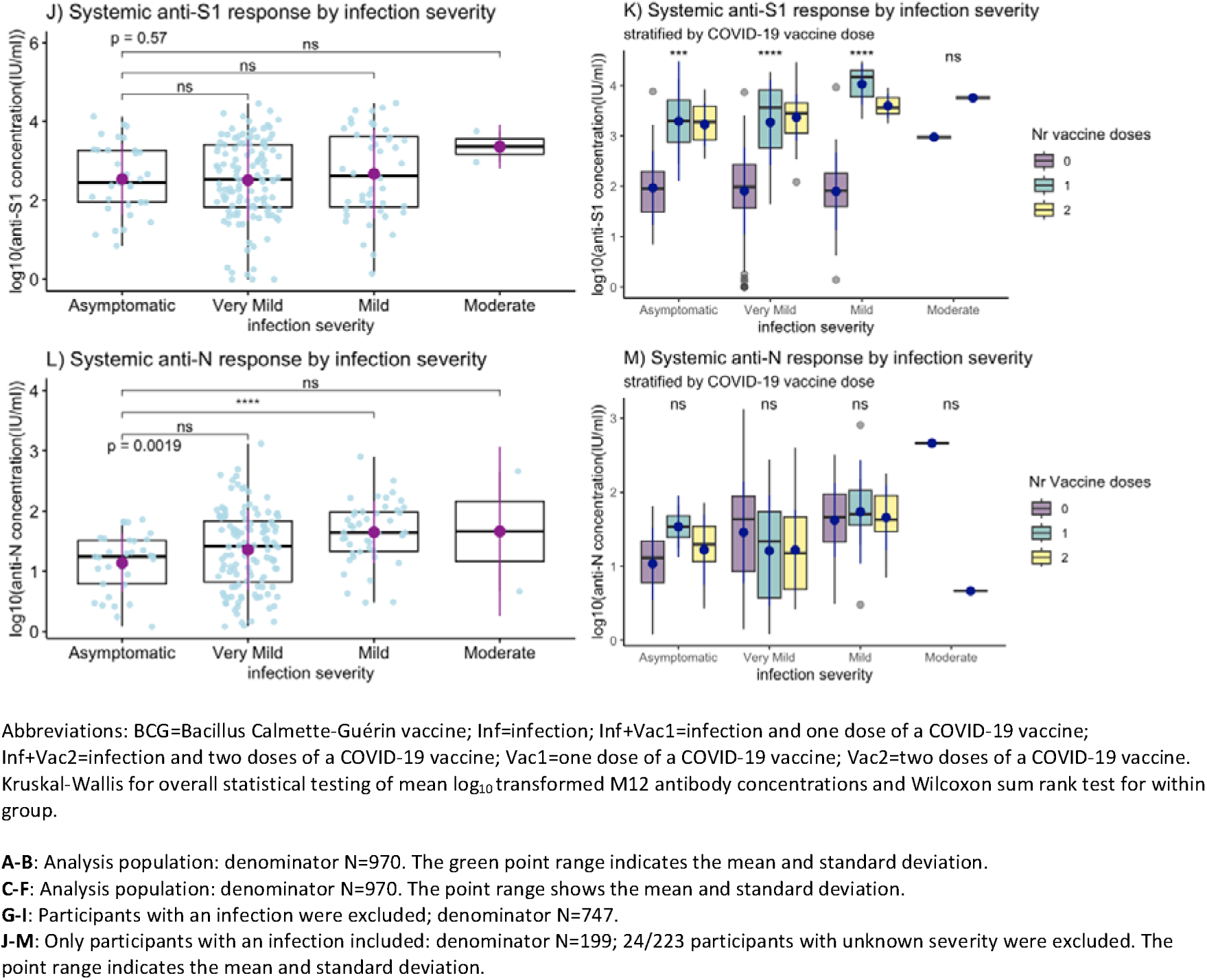
M12 anti-S1 and anti-N log_10_ concentrations by number and type of immune events.

**Table 2:**
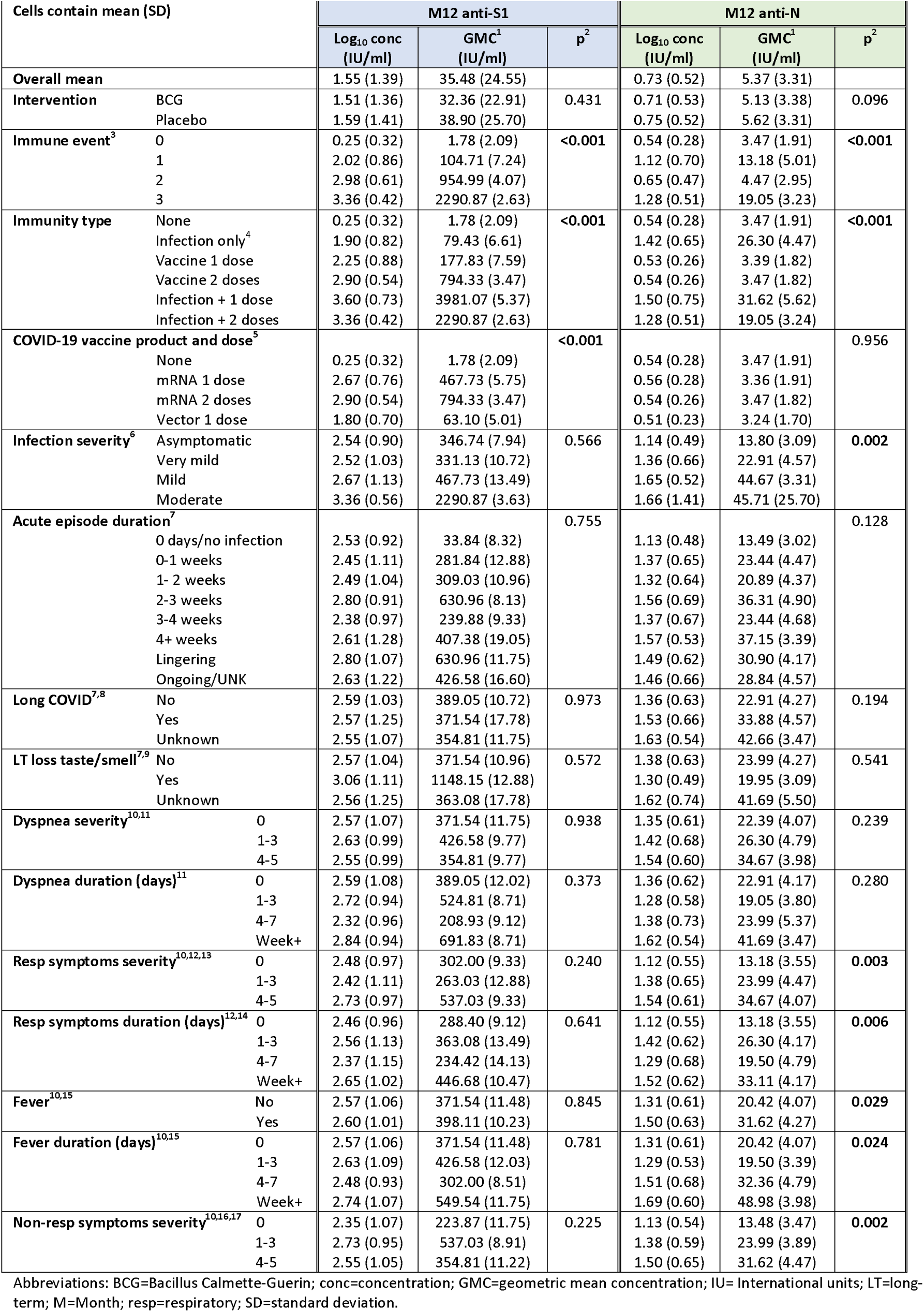
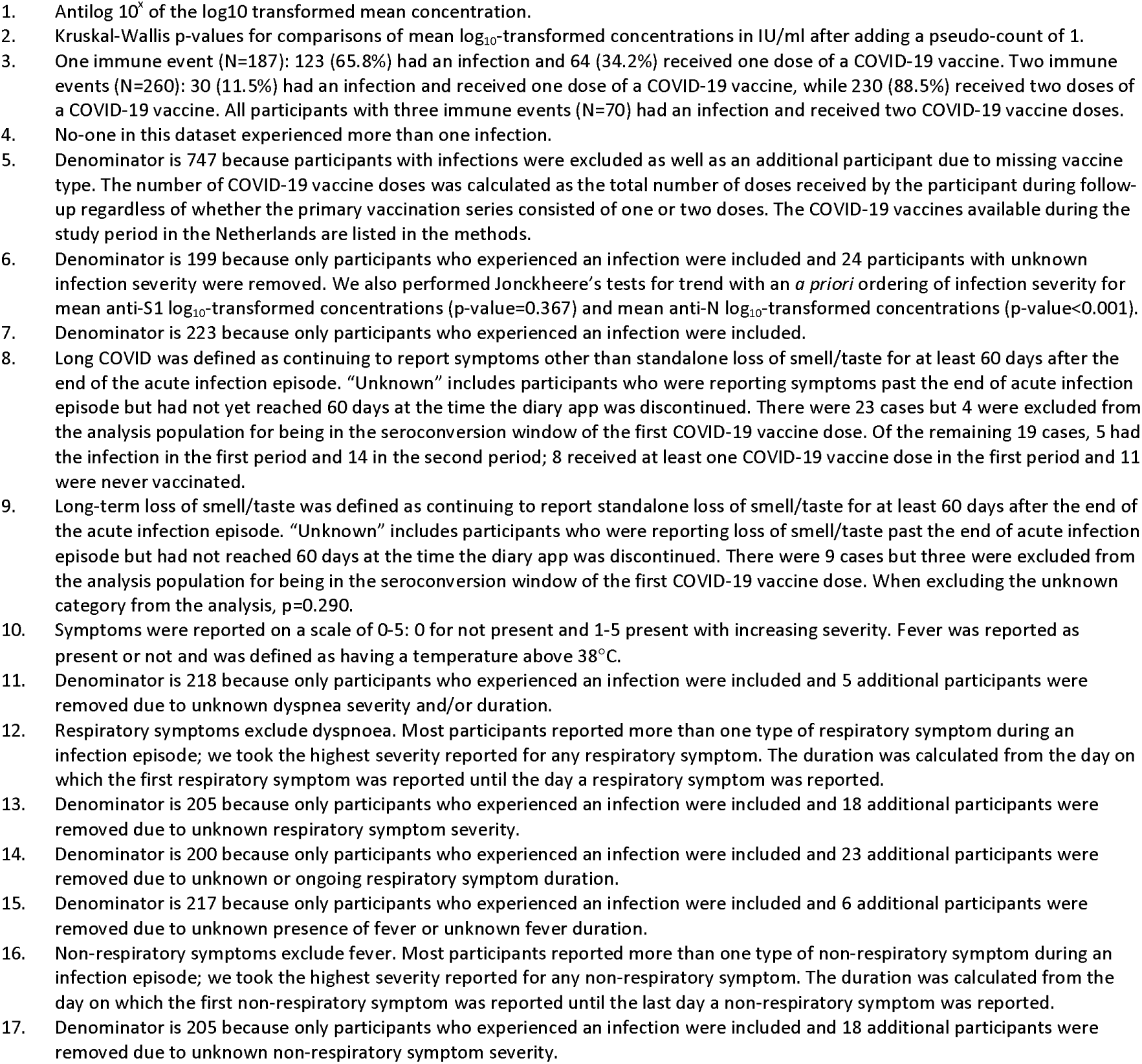
M12 anti-S1 and anti-N geometric mean concentrations by numbers and types of immune events.

### M12 anti-S1 and anti-N IgG GMCs by COVID-19 vaccinations

Of the 394/970 (40.6%) participants who received COVID-19 vaccination during follow-up, 94/970 (9.7%) received one dose and 300/970 (30.9%) received two doses. The majority (355/394; 90.1%) received an mRNA vaccine (305 Comirnaty, 49 Spikevax, and one an experimental vaccine by CureVac N.V. in a clinical trial setting), 36/394 (9.1%) received a viral vector vaccine (32 Vaxzevria and 4 Jcovden), and for three (0.8%) information on the vaccine type was missing (Table 1B). It should be noted that all participants who received two vaccine doses received mRNA vaccines, while those who received one dose received either a first dose of a mRNA vaccine (58.5%), a first or only dose of a vector vaccine (38.3%), or unknown vaccine type (3.2%). When excluding all participants who experienced an infection, M12 anti-S1 GMCs were significantly higher in participants who had received one dose of a mRNA vaccine (467.7 IU/ml; SD=5.8) compared to one dose of a viral vector vaccine (63.1 IU/ml; SD=5.0; p<0.001; Table 2, Figures 2G-I). Two doses of a mRNA vaccine resulted in a higher anti-S1 GMC than one (Table 2). M12 anti-S1 concentrations were higher in the Spikevax compared to the Comirnaty group; 562.3 IU/ml (SD=5.6) and 120.2 IU/ml (SD=4.6) for a single dose of Spikevax and Comirnaty, respectively (p=0.165), and 1230.2 IU/ml (SD=2.1) and 794.33 IU/ml (SD=3.5) for two doses of Spikevax and Comirnaty, respectively (p=0.583; Table S2). M12 anti-S1 GMCs were highest in participants who had an infection and also received one or two COVID-19 vaccinations: 3981.07 IU/ml (SD=5.37) and 2290.87 IU/ml (SD=2.63), respectively (Table 2).

### M12 anti-S1 and anti-N IgG GMCs by SARS-CoV-2 infections

A total of 223 first infections occurred in the analysis population during follow-up: 33 (14.8%) were asymptomatic, 120 (53.8%) very mild, 44 (19.7%) mild, 2 (0.90%) moderate, and 24 (10.8%) of unknown severity. Among those who experienced an infection during follow-up, 30/223 (13.5%) received one COVID-19 vaccine dose and 70/223 (31.4%) received two doses. The M12 anti-S1 GMCs did not statistically significantly differ by overall infection severity when analysed as an ordinal variable (p=0.566), and this remained the case after stratifying by the number of COVID-19 vaccine doses received (Table 2, Figures 2J-K). In contrast, the M12 anti-N GMCs significantly increased with increasing overall infection severity: 13.8 IU/ml (SD=3.1) for asymptomatic, 22.9 IU/ml (SD=4.6) for very mild, 44.7 IU/ml (SD=3.3) for mild, and 45.7 IU/ml (SD=25.7) for moderate infections (p=0.002; Table 2, Figures 2L-M). The M12 anti-S1 and anti-N GMCs were not associated with increasing acute episode duration, Long COVID or long-term loss of smell/taste (Table 2, Figures S5A-F). M12 anti-S1 GMCs were not associated with the severity or duration of specific individual symptoms (analysed as ordinal variables) either. In contrast, M12 anti-N GMCs were higher with higher severity and longer duration of respiratory symptoms excluding dyspnoea, with the presence and duration of fever, and with the severity of non-respiratory symptoms other than fever (Table 2, Figures S5G-T). Of note, the severity and duration of dyspnoea were not associated with either M12 anti-S1 or M12 anti-N GMCs (Table 2, Figure S5G-J).

The M12 anti-S1 and anti-N GMCs of participant who experienced an infection were correlated at the individual level: participants with a higher M12 anti-S1 GMC also had a higher M12 anti-N GMC (Figure S6). This correlation was most pronounced for those who never received a COVID-19 vaccine during follow-up (Spearman’s ρ=0.69; p<0.001) and weakened in participants who had received one (ρ=0.54; p=0.003) or two vaccinations (ρ=0.32; p=0.006) that boosted anti-S1 GMCs only.

### Linear regression models of M12 anti-S1 and anti-N concentrations

In univariable models including the entire analysis population (N=970), the number of immune events, immune event type, overall infection severity (analysed as an indicator variable), and work-related exposure risk were statistically significantly associated with both M12 anti-S1 and anti-N log_10_ concentrations (Table S3A). Vaccine doses and types (combined into one variable) were only associated with M12 anti-S1 log_10_ concentration, while hypertensive medication use and history of pulmonary disease other than asthma and hay fever were only associated with M12 anti-N log_10_ concentrations. BCG/placebo vaccination at baseline, age, sex, smoking, history of BCG vaccination, influenza vaccination during follow-up, recent respiratory tract infections, and the comorbidities that were not already mentioned were not associated with M12 anti-S1 or anti-N log_10_ concentrations (Table S3A). In univariable subgroup analyses limited to participants who experienced an infection (n=223), overall infection severity, overall acute episode duration, and specific symptom severity and duration, with the exception of dyspnoea, were significantly associated with M12 anti-N log_10_ concentrations (all as indicator variables; Table S3B). Long COVID and long-term loss of smell/taste were not associated with M12 anti-S1 or anti-N log_10_ concentrations in individuals who had experienced an infection.

The final multivariable models with M12 anti-S1 or anti-N log_10_ concentrations as the outcomes and BCG/placebo vaccination at baseline, age, and sex forced into the models are shown in Table 3. Age, vaccine type plus dose (indicator variable), and overall infection severity (indicator variable) were statistically significantly associated with M12 anti-S1 log_10_ concentration. Sex and overall infection severity (indicator variable) were associated with M12 anti-N log_10_ concentration. Current use of hypertensive medication was associated with M12 anti-N log_10_ concentration in univariable analysis but was no longer significant in the multivariable model. Sensitivity analyses adding more covariates (Figure S7), or reducing the seroconversion windows to 7 and 0 days (Table S4), produced similar results with two exceptions. The positive associations between M12 anti-S1 log_10_ concentration and mild or moderate infection severity (compared to no infection) and having received one dose of a mRNA or vector vaccine (compared to no vaccination) changed in magnitude, but remained statistically significant, when the seroconversion period was reduced to 7 or 0 days (Table S4; Supplementary Results).

**Table 3:**
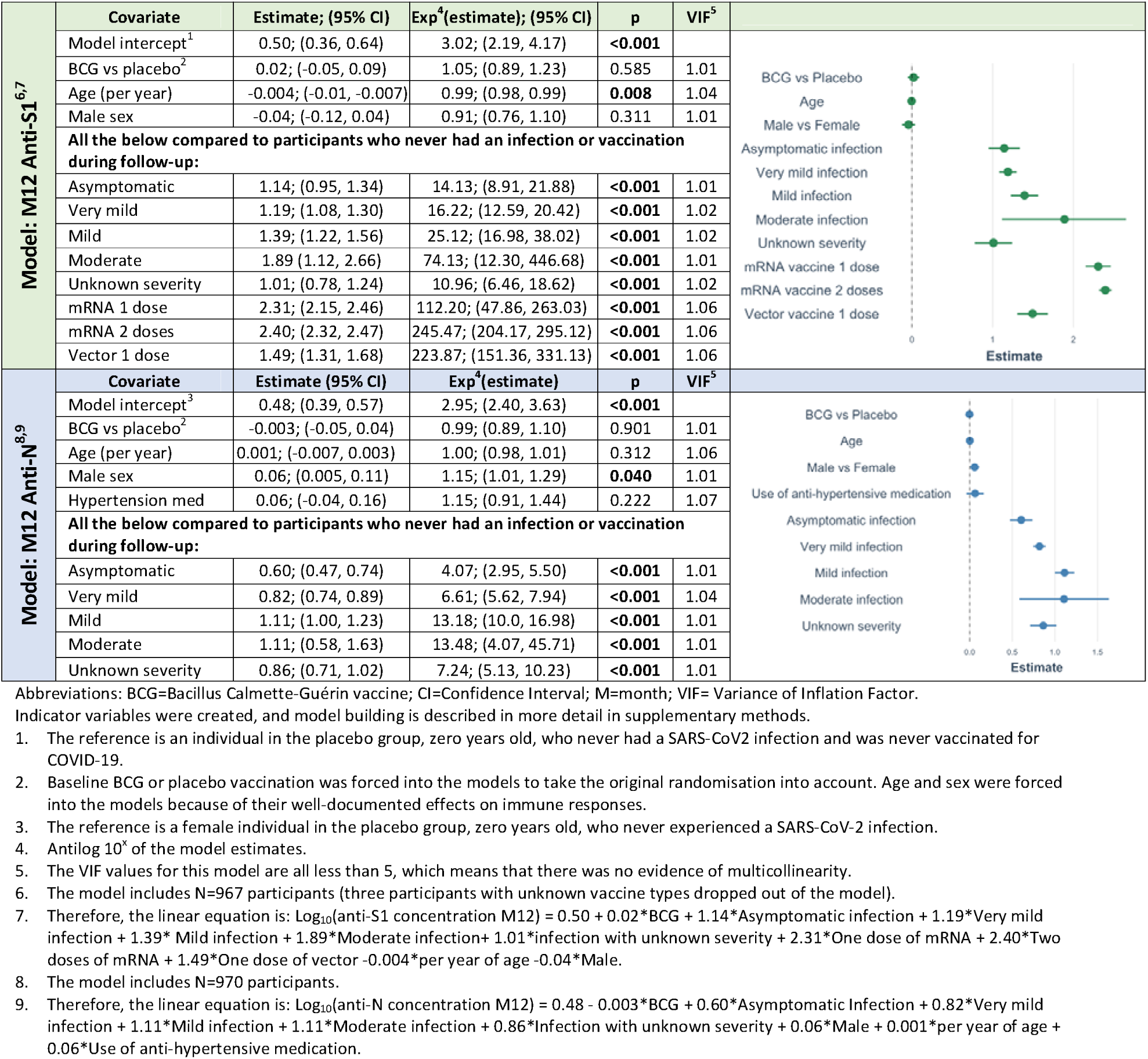
Multivariable models with M12 anti-S1 and anti-N log_10_ concentrations as outcome.

| Model: M12 Anti-S1<sup>6,7</sup> | Covariate | Estimate; (95% CI) | Exp<sup>4</sup>(estimate); (95% CI) | p | VIF<sup>5</sup> |
| --- | --- | --- | --- | --- | --- |
|  | Model intercept<sup>1</sup> | 0.50; (0.36, 0.64) | 3.02; (2.19, 4.17) | <0.001 |  |
|  | BCG vs placebo<sup>2</sup> | 0.02; (-0.05, 0.09) | 1.05; (0.89, 1.23) | 0.585 | 1.01 |
|  | Age (per year) | -0.004; (-0.01, -0.007) | 0.99; (0.98, 0.99) | **0.008** | 1.04 |
|  | Male sex | -0.04; (-0.12, 0.04) | 0.91; (0.76, 1.10) | 0.311 | 1.01 |
|  | All the below compared to participants who never had an infection or vaccination during follow-up: |  |  |  |  |
|  | Asymptomatic | 1.14; (0.95, 1.34) | 14.13; (8.91, 21.88) | <0.001 | 1.01 |
|  | Very mild | 1.19; (1.08, 1.30) | 16.22; (12.59, 20.42) | <0.001 | 1.02 |
|  | Mild | 1.39; (1.22, 1.56) | 25.12; (16.98, 38.02) | <0.001 | 1.02 |
|  | Moderate | 1.89 (1.12, 2.66) | 74.13; (12.30, 446.68) | <0.001 | 1.01 |
|  | Unknown severity | 1.01; (0.78, 1.24) | 10.96; (6.46, 18.62) | <0.001 | 1.02 |
|  | mRNA 1 dose | 2.31; (2.15, 2.46) | 112.20; (47.86, 263.03) | <0.001 | 1.06 |
|  | mRNA 2 doses | 2.40; (2.32, 2.47) | 245.47; (204.17, 295.12) | <0.001 | 1.06 |
|  | Vector 1 dose | 1.49; (1.31, 1.68) | 223.87; (151.36, 331.13) | <0.001 | 1.06 |
| Model: M12 Anti-N<sup>8,9</sup> | Covariate | Estimate (95% CI) | Exp<sup>4</sup>(estimate) | p | VIF<sup>5</sup> |
|  | Model intercept<sup>3</sup> | 0.48; (0.39, 0.57) | 2.95; (2.40, 3.63) | <0.001 |  |
|  | BCG vs placebo<sup>2</sup> | -0.003; (-0.05, 0.04) | 0.99; (0.89, 1.10) | 0.901 | 1.01 |
|  | Age (per year) | 0.001; (-0.007, 0.003) | 1.00; (0.98, 1.01) | 0.312 | 1.06 |
|  | Male sex | 0.06; (0.005, 0.11) | 1.15; (1.01, 1.29) | **0.040** | 1.01 |
|  | Hypertension med | 0.06; (-0.04, 0.16) | 1.15; (0.91, 1.44) | 0.222 | 1.07 |
|  | All the below compared to participants who never had an infection or vaccination during follow-up: |  |  |  |  |
|  | Asymptomatic | 0.60; (0.47, 0.74) | 4.07; (2.95, 5.50) | <0.001 | 1.01 |
|  | Very mild | 0.82; (0.74, 0.89) | 6.61; (5.62, 7.94) | <0.001 | 1.04 |
|  | Mild | 1.11; (1.00, 1.23) | 13.18; (10.0, 16.98) | <0.001 | 1.01 |
|  | Moderate | 1.11; (0.58, 1.63) | 13.48; (4.07, 45.71) | <0.001 | 1.01 |
|  | Unknown severity | 0.86; (0.71, 1.02) | 7.24; (5.13, 10.23) | <0.001 | 1.01 |

Abbreviations: BCG=Bacillus Calmette-Guérin vaccine; CI=Confidence Interval; M=month; VIF= Variance of Inflation Factor.
Indicator variables were created, and model building is described in more detail in supplementary methods.

## Discussion

SARS-CoV-2 anti-S1 and anti-N GMCs approximately one year after the start of the Dutch SARS-CoV-2 epidemic were associated with the number of immune events, and overall infection severity when an infection took place, but not with having had or having Long COVID or long-term loss of smell/taste. Anti-S1 GMCs were also associated with COVID-19 vaccination, and anti-N GMCs with specific symptom severity and duration except for dyspnoea. Anti-S1 GMCs were higher after one vaccination than one infection, and after one mRNA than one vector vaccination, and highest after a combination of infection and vaccination(s).

Our finding of higher anti-S1 GMCs with an increasing number of immune events, and after vaccination with mRNA compared to vector vaccines, is in agreement with the published literature [23–25]. However, published reports for infection versus vaccination are inconsistent [26]. Important variables in that context are the severity and duration of infections, waning antibody concentrations over time, and the timing of boosting events. We were able to investigate the former, but not the latter. We did not detect a downward trend of antibody GMCs by time period between the last immune event and M12 sampling, but we could not evaluate antibody concentrations longitudinally for individual participants due to the low number of immune events in the first seroconversion period. The COVID-19 vaccination campaign in the Netherlands began eight months after the start of the study and SARS-CoV-2 infections took place throughout follow-up. Our finding of higher anti-S1 GMC after one vaccination than after one infection could therefore be explained by the timing of immune events and subsequent waning antibody concentrations over time. However, the follow-up time was short with little opportunity for waning, and our findings were robust after applying seroconversion windows of 14, 7 or 0 days. Another potential explanation is lower antigen exposure during a mild infection than after vaccination. Participants with hybrid immunity due to infection plus vaccination(s) had higher anti-S1 GMCs than those with infection or vaccination (one or two doses) immunity only. This is also in agreement with the published literature [25, 27, 28]. Studies have shown that within the hybrid immunity group, concentrations depend on the type(s) of vaccine(s) used, the number of doses, and the severity of the infection [28–30].

As we had already shown earlier, BCG vaccination administered in the beginning of the study did not influence SARS-CoV-2 specific antibody concentrations approximately one year later. This is in agreement with most other BCG trials conducted during the COVID-19 epidemic failing to demonstrate benefits of BCG vaccination on COVID-19-related endpoints [18, 19, 31–33].

Overall infection severity was associated with M12 anti-S1 (when analysed as an indicator variable but not as an ordinal variable) and anti-N GMCs (when analysed as an indicator or ordinal variable), and individual symptom severity and durations with M12 anti-N GMCs only. However, the latter associations did show non-statistical trends in the same direction for M12 anti-S1 GMCs. The somewhat inconsistent findings for higher M12 anti-S1 GMCs with increasing infection severity could be explained by the fact that this increase was non-linear, with GMCs starting out higher than anti-N GMCs (likely because the host immune system is highly exposed to the viral S1-protein and less so the N-protein)[3] and reaching a plateau more easily. Several other studies have reported increasing anti-S1 and anti-N antibody concentrations with increasing infection severity [1, 13, 34], but some studies did not find this association [35, 36]. Many of these studies, however, compared WHO-defined mild, moderate and severe infections (with the latter two requiring hospitalisation). Our study was conducted in a generally healthy population below the age of 65, and the detailed and comprehensive diary data allowed us to also include asymptomatic infections and to subdivide the WHO-defined mild category into very mild and mild subcategories. Even within this range of asymptomatic to mild infections differences in impact on antibody responses were measurable.

Our analyses did not reveal associations between M12 antibody concentrations and having had or having Long COVID or long-term loss of smell/taste, but the analysis population included only 19 Long COVID cases. Several other studies reported weaker SARS-CoV-2-specific antibody responses following an infection in Long COVID cases [37–39]. However, one study reported similar anti-S1 and anti-N concentrations between fully recovered and Long COVID patients after infection but aberrant immune responses in Long COVID patients following COVID-19 vaccination [40]. The potential associations between immune responses and development of Long COVID requires further study.

The main strengths of the study were that we captured the entire first year of the Dutch COVID-19 epidemic and had detailed and comprehensive data, allowing us to identify asymptomatic infections and to subdivide WHO mild infections into very mild and mild. A main limitation was that we could not account for antibody waning at the individual participant level, but we think that follow-up time after most immune events was too short for waning to have influenced results significantly. Other limitations include the use of two different blood sampling methods (venepuncture and fingerprick sampling) and the suboptimal sensitivity of the anti-N assay for detecting asymptomatic infections (67%) [9]. We may therefore have missed some asymptomatic infections and overestimated the M12 anti-N GMC in the asymptomatic group.

### Conclusions

Our study confirmed that anti-S1 and anti-N GMCs were associated with overall infection severity, but additionally showed that this was the case even in infections ranging from asymptomatic to mild in an otherwise healthy population. Anti-S1 GMCs were also associated with numbers and types of COVID-19 vaccinations, with the combination of infection and vaccination(s) eliciting the greatest response. Anti-S1 and anti-N GMCs were not associated with having had of having Long COVID or long-term loss of smell/taste.

## Data Availability

Individual participant data that underlie the results reported in this article will be made available after deidentification to investigators whose proposed use of the data has been approved by an independent review committee up to 5 years following publication. The study protocol will be available to anyone during this same time frame. Information regarding submitting proposals and accessing data may be found on https://dataverse.nl/.

## Acknowledgments

We thank the healthcare workers for their participation, and colleagues in the UMC Utrecht and participating hospitals and laboratories who implemented the BCG-Corona trial during the Dutch COVID-19 epidemic.

## Author contributions

JW, TD, MB and MN designed and obtained funding for the study. TD and JW supervised the clinical data collection, and JW supervised the analyses and paper-writing. GS, RB, and FK conducted the antibody testing and MJ and LV provided additional immunological expertise. JC contributed to the clinical data collection, conducted the analyses, and wrote the first draft of the paper. All authors read and approved the final version. JC and JW have full access to all data and take responsibility for the integrity of the data and the accuracy of the data analyses.

