## Supplementary Material for "Systemic SARS-CoV-2-specific antibody responses to infection and to COVID-19 and BCG vaccination"

**SUPPLEMENTARY METHODS**

The sample collection protocol, the antibody measurement procedure, and outcome definitions have been previously described in detail [1].

**Inclusion and exclusion criteria**

Participants were healthcare workers, including doctors, nurses, paramedics, and support staff. They were 18 or older and had to be in possession of a smartphone. Participants were also expected to be working in direct contact with SARS-CoV-2 infected patients. The primary exclusion criteria were known allergy to BCG, active or latent *Mycobacterium tuberculosis* infection (as judged by the local Principal Investigator in each hospital), any other active infection, immunocompromised state, malignancy or lymphoma in the past two years, current or planned pregnancy, any vaccination in the past 4 weeks, having a hospital employment contract of less than 22 hours per week, or expected work absence of at least 4 weeks.

**Sample collection**

About half of the participants were recruited in the three core hospitals (University Medical Center (UMC) Utrecht, Radboud UMC, and Leiden UMC) and invited to participate in sampling in their own hospital at about 3 months (M3; June 2020) and 12 months (M12; April 2021) after study vaccination. One serum aliquot per participant was transported to the Center for Immunology of Infections and Vaccines at the National Institute for Public Health and the Environment (RIVM in Dutch) in Bilthoven, the Netherlands, for antibody testing.

The remaining participants were recruited in six other hospitals (Noordwest Ziekenhuis Alkmaar, Haga Ziekenhuis Den Haag, Canisius-Wilhelmina Ziekenhuis Nijmegen, Sint Maartenskliniek Nijmegen, Jeroen Bosch Ziekenhuis Den Bosch, and Erasmus Medisch Centrum Rotterdam), and were invited to fingerprick sampling at home. Participants from core hospitals who could not attend an in-hospital sampling visit were also asked to conduct finger-prick sampling at home. The implementation of each at-home sampling round took several weeks; the first round took place from 9 Oct 2020 until 18 Dec 2020, and the second round from 14 April 2021 until 9 June 2021. The diary app was discontinued on 27 March 2021. We collected data on COVID-19-like symptoms, SARS-CoV-2 tests, and COVID-19 vaccinations between 27 March 2021 (diary app cessation) and the participant’s final sampling date via Formdesk (Innovero Software Solutions BV, Wassenaar, Netherlands) and email.

**Diary completeness**

The app completion percentage was calculated for the period between vaccination and app discontinuation on 27 March 2021, and the total follow-up time as the number of days between vaccination and the M12 sampling date. The app completion percentage was calculated as the number of completed app entries (the number of days for which the participant completed the diary, regardless of whether the s/he formally withdrew from the study) divided by the expected number of app entries (the number of days between randomisation and diary app cessation on 27 March 2021). Participants were considered to have adequate follow-up data if the app completion percentage was at least 80%.

**Seroconversion window**

The seroconversion window was defined as 14 days prior to the participant’s M12 sampling date until the date of sampling. We used 14 days because previous studies have shown that the median systemic IgG seroconversion time is around 14 days for hospitalised patients [2], with more severe cases seroconverting at a faster rate than mild cases [3]. Furthermore, in mild cases, antibodies are reliably detected between 10-17 days after symptoms onset [2–4]. We conducted sensitivity analyses with seroconversion windows of 0 and 7 days instead of 14 days (Table S4). The numbers of participants that were excluded because they had an immune event within the seroconversion window are shown in the footnotes of Table S4.

**Covariates**

*Work-related SARS-CoV-2 exposure risk*

We combined several baseline workplace characteristics (job function and department, percent of work hours with direct patient contact, and planned work on a COVID-ward) into one workplace exposure risk variable, categorised as high, medium, or low risk as follows:

|  |  | Planned work in COVID-ward | | |
| --- | --- | --- | --- | --- |
| % work hours with patient contact |  | **Yes** | **Unknown** | **No** |
|  | **0-25** | Medium | Medium | Low |
|  | **26-50** | High | Medium | Low |
|  | **51-75** | High | High | Medium |
|  | **75+** | High | High | Medium |

*Positive SARS-CoV-2 infection*

A positive SARS-CoV-2 infection was identified through self-reporting of a positive SARS-CoV-2 test of any type or seroconversion as described in the methods of the manuscript. Four participants reported a second infection during the one year follow-up. These participants were excluded from the analysis population because the subgroup was too small to generate meaningful results.

*Immune events, and vaccination types and doses*

These covariates are described in the methods of the manuscript.

*Overall infection severity and severity of individual symptoms*

The following respiratory and non-respiratory symptoms could be reported in the diary app:

Respiratory symptoms (all reported on a scale of 0-5, with 0 meaning not present):

- Nose cold (*Neusverkouden*)
- Sore throat (*Keelpijn*)
- Cough (*Hoesten*)
- Dyspnea/shortness of breath (*Kortademig*)
- Loss of smell/taste (*Reuk en/of smaakverlies*)

Non-respiratory symptoms (all reported on a scale of 0-5 except fever):

- Fever (*Koorts*), defined as a temperature of 38 °C or above
- Cold shivers (*Koude rillingen*)
- Muscle pain (*Spierpijn*)
- Fatigue (*Vermoeidheid*)
- Headache (*Hoofdpijn*)
- Diarrhoea (*Diarree*)

We used the World Health Organization (WHO) definitions for infection severity [5,6], and further subcategorised WHO mild category into very mild and mild subcategories. The definitions of episode severity are described below. Chronic symptoms that were consistently reported by participant without a clear link to a test date or episode of respiratory symptoms were ignored.

- *WHO moderate:* Participant had clinical signs of pneumonia and hospitalisation due to SARS-CoV-2 but did not require high flow oxygen therapy.
- *WHO mild:* Participants experienced symptoms due to SARS-CoV-2 but were not hospitalised and did not have evidence of pneumonia or hypoxia.
  - *Subcategory mild:* Participant reported a fever (temperature 38+) for more than one week AND/OR dyspnoea reaching level 4 or 5 for more than one week AND/OR any other respiratory symptoms with at least one symptom other than loss of smell/taste reaching level 4 or 5 for more than one week AND/OR a total symptomatic episode lasted for more than 28 days.
  - *Subcategory very mild:* Participant reported symptoms during infection episode that did not reach the level of subcategory mild as described above.
- *WHO asymptomatic:* Participant did not experience any symptoms during relevant period, infection episode was detected via SARS-CoV-2 diagnostic testing or serology.

When it was impossible to draw conclusions about episode severity from the available data, the episode severity was coded as unknown.

*Overall acute episode duration and duration of individual symptoms*

The overall acute episode duration was calculated as the number of consecutive days during which a participant reported symptoms, excluding Long COVID or long-term loss of smell/taste. Similar definitions were used for each individual symptom. Long COVID (lingering symptoms other than standalone loss of smell/taste) and long-term loss of smell/taste were defined as continuing to report symptoms for at least 60 consecutive days after the end of the acute infection episode [7].

**Regression model building**

Associations between covariates and log_10_-transformed M12 anti-S1 and anti-N concentrations were assessed using univariable and multivariable linear regression models for the full analysis population (Table 3, Table S3A). First, all covariates were assessed in univariable models, and those with p<0.05 were considered for inclusion in multivariable models. However, we decided to exclude:

- work-related SARS-CoV-2 exposure risk because it determines someone’s likelihood of becoming infected rather than the immune response to infection.
- past TB test results because the ‘not done’ and ‘unknown’ categories were the only categories that were statistically significant in the anti-S1 univariable model.
- having received an influenza vaccination in the winter of 2019/2020 because the ‘missing’ category was the only statistically significant category in the anti-S1 univariable model.
- history of other pulmonary disease (other than hay fever or asthma) because of low prevalence.

We added these variables to the final model in sensitivity analyses (Figure S7).

In addition, the use of anti-hypertensive medication was kept in the multivariable anti-N model despite it no longer being significant because it has been consistently associated with SARS-CoV-2 infection risk and severity. There may be a biological reason for this: patients with high blood pressure have upregulated angiotensin-converting enzyme 2 (ACE2), which is the entry receptor for SARS-CoV-2 [8,9].

Various covariates were strongly correlated. Severity of infection was strongly correlated with having an infection (a prerequisite) and with duration of acute infection episode (included in the severity definition). We therefore created a new variable called ‘overall infection severity’, including a ‘no infection’ category, and did not add acute episode duration to the final multivariable models (we did add it as a sensitivity analysis as shown in Figure S7). The severities and durations of individual symptoms were correlated with each other (see correlation matrix below) and with overall infection severity and were therefore only assessed in univariable models.

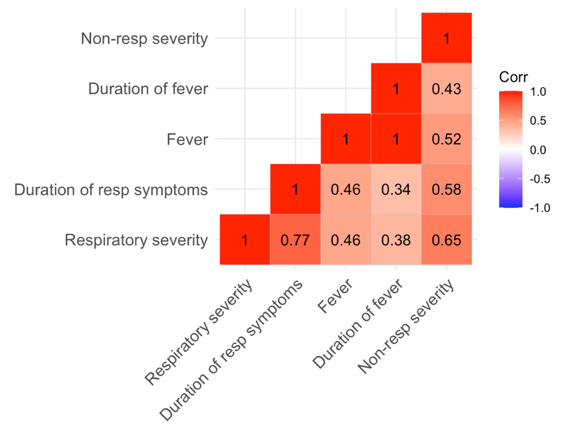

The COVID-19 vaccine type was correlated with the number of doses received due to vaccination recommendations and roll-out procedures: most healthcare workers qualified for mRNA vaccination early on during roll-out, whereas the vector vaccines were rolled out later and the primary schedule of vector vaccine Jcovden consisted of only one dose. Therefore, by the end of our follow-up period, most participants who received a mRNA vaccine reported having received two doses of that vaccine, whereas all participants that received a vector vaccine only reported one dose. We therefore created a new COVID-19 vaccination variable with the categories not vaccinated, mRNA vaccine 1 dose, mRNA vaccine 2 doses, and vector vaccine 1 dose.

In addition to the above-described univariable and multivariable analyses on the full analysis population, we also ran univariable models on a subgroup of participants who experienced an infection (Table S3B).

The final models adhered to the univariable and multivariable linear regression assumptions, and the variance inflation factor (VIF) values of the multivariable models were less than 5, indicating a lack of multicollinearity.

**SUPPLEMENTARY RESULTS**

**Sensitivity analyses results**

Sensitivity analyses adding more covariates to the final multivariable models produced similar results (Figure S7). Overall infection severity and overall acute episode duration could not be added due to multicollinearity.

Sensitivity analyses reducing the seroconversion windows from 14 to 7 or 0 days also produced similar results with two exceptions. The positive associations between M12 anti-S1 log_10_ concentration and mild or moderate infection severity (compared to no infection) increased when the seroconversion period was reduced, and having received one dose of a mRNA or vector vaccine (compared to no vaccination) decreased, but estimates were still statistically significant (Table S4). We explored reasons for the changes in the strengths of the associations. Only two moderate infections were included in the analysis population after applying a 14 day seroconversion window, and this increased to three infections when the window was reduced to 7 or 0 days. These numbers are too small to enable meaningful conclusions. The observed increase in the estimate for mild infections may be due to potential boosting by COVID-19 vaccination after experiencing an infection. Most of the participants in the 14-day seroconversion window were in that window because of a recent vaccination, not because of a recent infection.

[5]	World Health Organization. COVID-19 clinical management: living guidance, 25 January 2021. World Health Organization; 2021.

[6]	World Health Organisation. A minimal common outcome measure set for COVID-19 clinical research. Lancet Infect Dis 2020;20:e192–7. https://doi.org/10.1016/S1473-3099(20)30483-7.

[7]	Soriano JB, Murthy S, Marshall JC, Relan P, Diaz JV, WHO Clinical Case Definition Working Group on Post-COVID-19 Condition. A clinical case definition of post-COVID-19 condition by a Delphi consensus. Lancet Infect Dis 2022;22:e102–7. https://doi.org/10.1016/S1473-3099(21)00703-9.

[8]	Úri K, Fagyas M, Kertész A, Borbély A, Jenei C, Bene O, et al. Circulating ACE2 activity correlates with cardiovascular disease development. J Renin-Angiotensin-Aldosterone Syst JRAAS 2016;17:1470320316668435. https://doi.org/10.1177/1470320316668435.

[9]	Li XC, Zhang J, Zhuo JL. The vasoprotective axes of the renin-angiotensin system: Physiological relevance and therapeutic implications in cardiovascular, hypertensive and kidney diseases. Pharmacol Res 2017;125:21–38. https://doi.org/10.1016/j.phrs.2017.06.005.

**SUPPLEMENTARY TABLES AND FIGURES**

**Figure S1: Distribution of log_10_ concentrations of A) anti-S1 IgG and B) anti-N IgG at M12**

**A)**

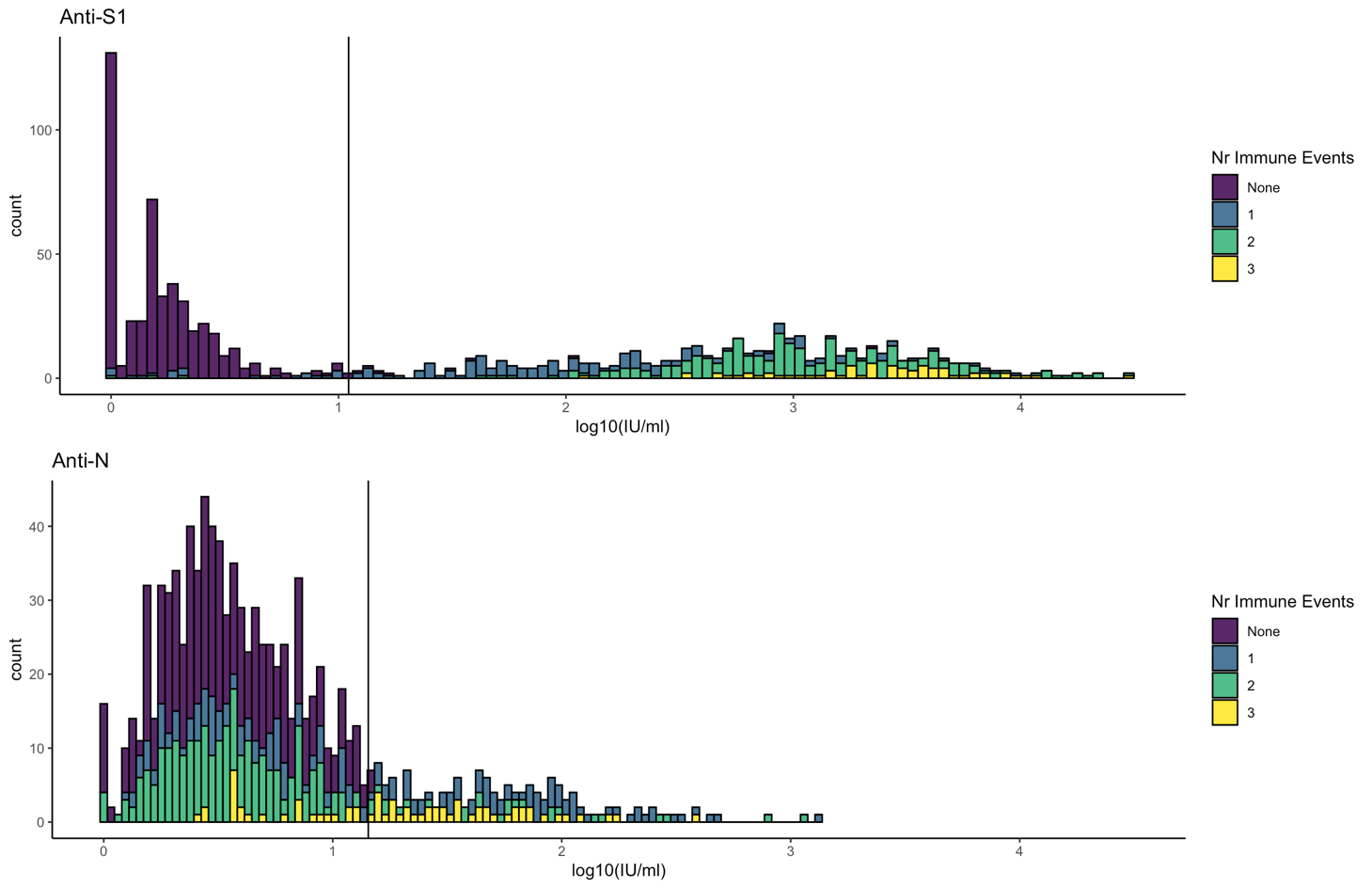

**B)**

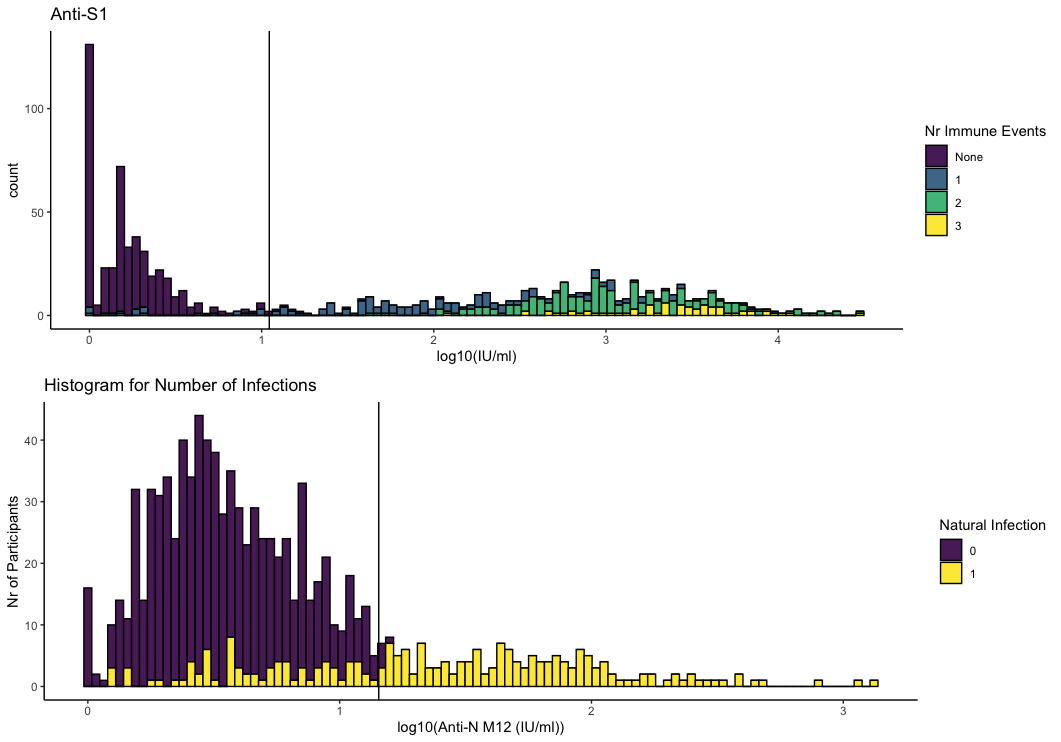

Systemic antibody concentrations were log_10_ transformed (after pseudocount of +1). The vertical lines in the panels marks the thresholds for anti-S1 and anti-N seropositivity, respectively. An immune event is either one SARS-CoV-2 infection or one dose of a COVID-19 vaccine.

**Figure S2:	Timelines of first COVID-19 vaccination (A-C) or SARS-CoV-2 infection (D-F) for seroconversion windows of 14, 7 or 0 days**

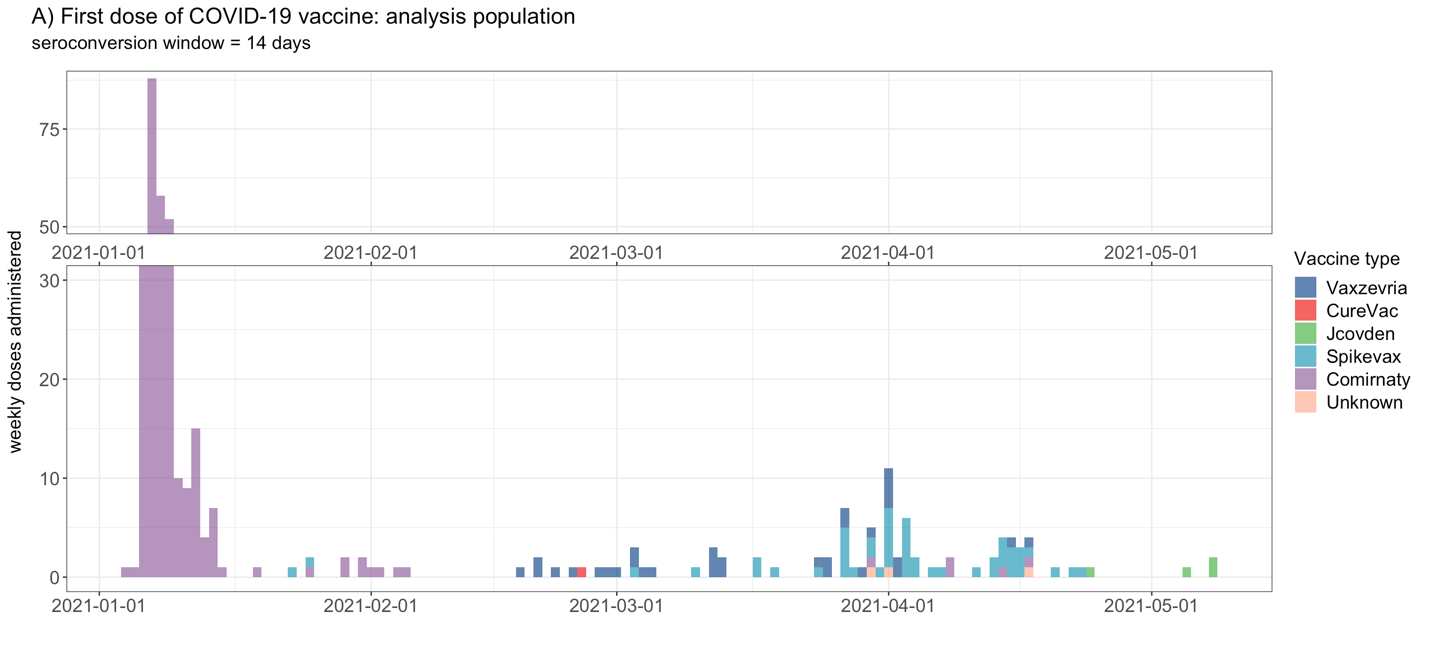

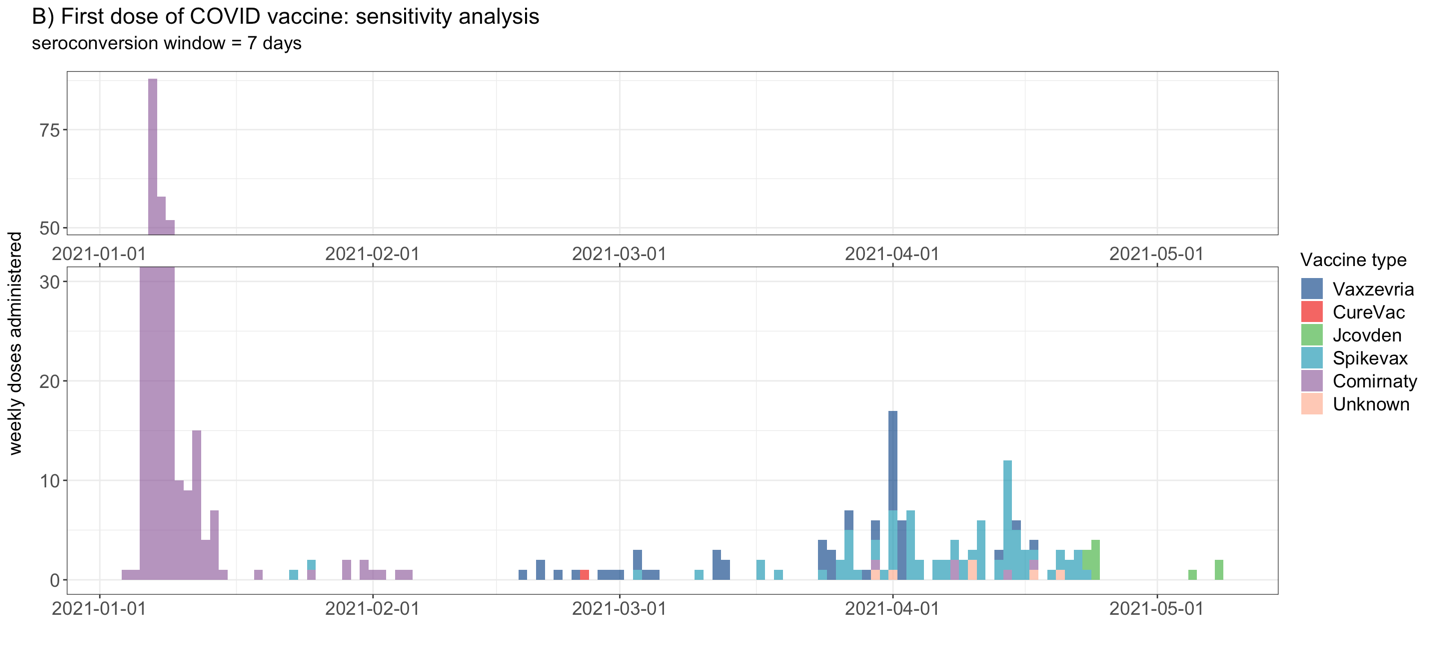

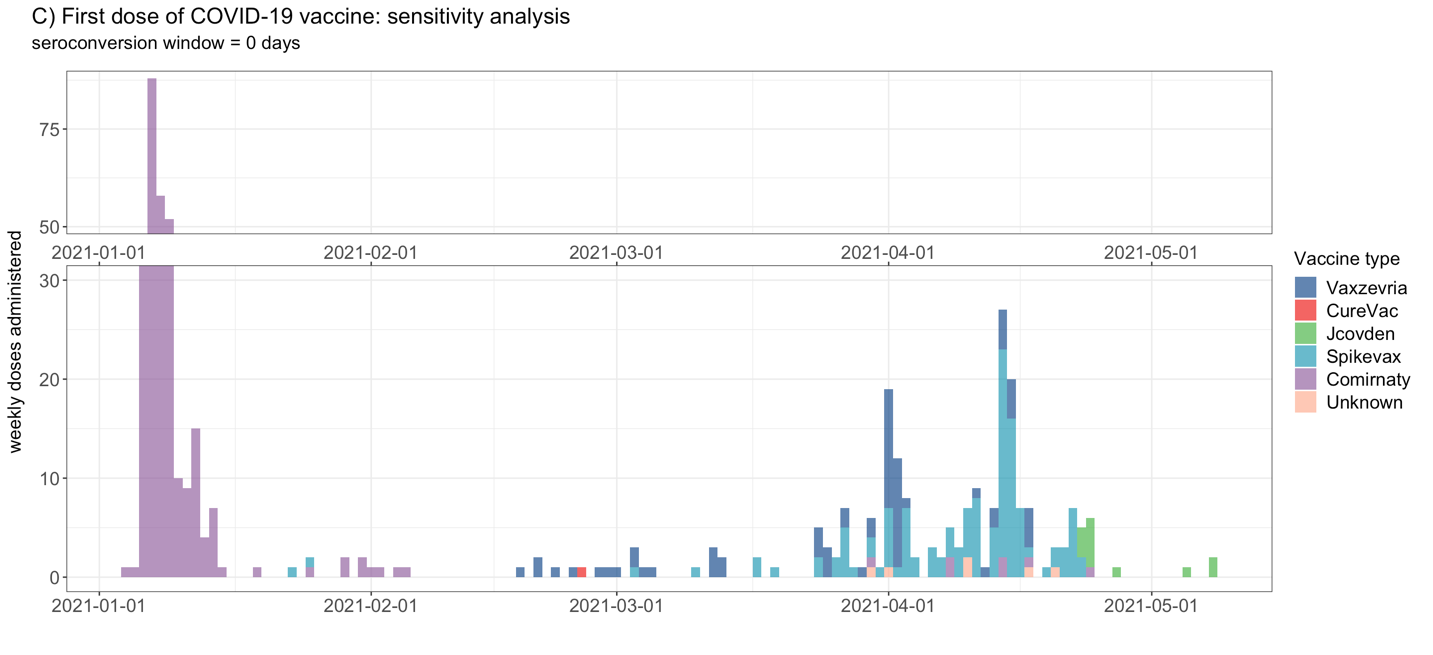

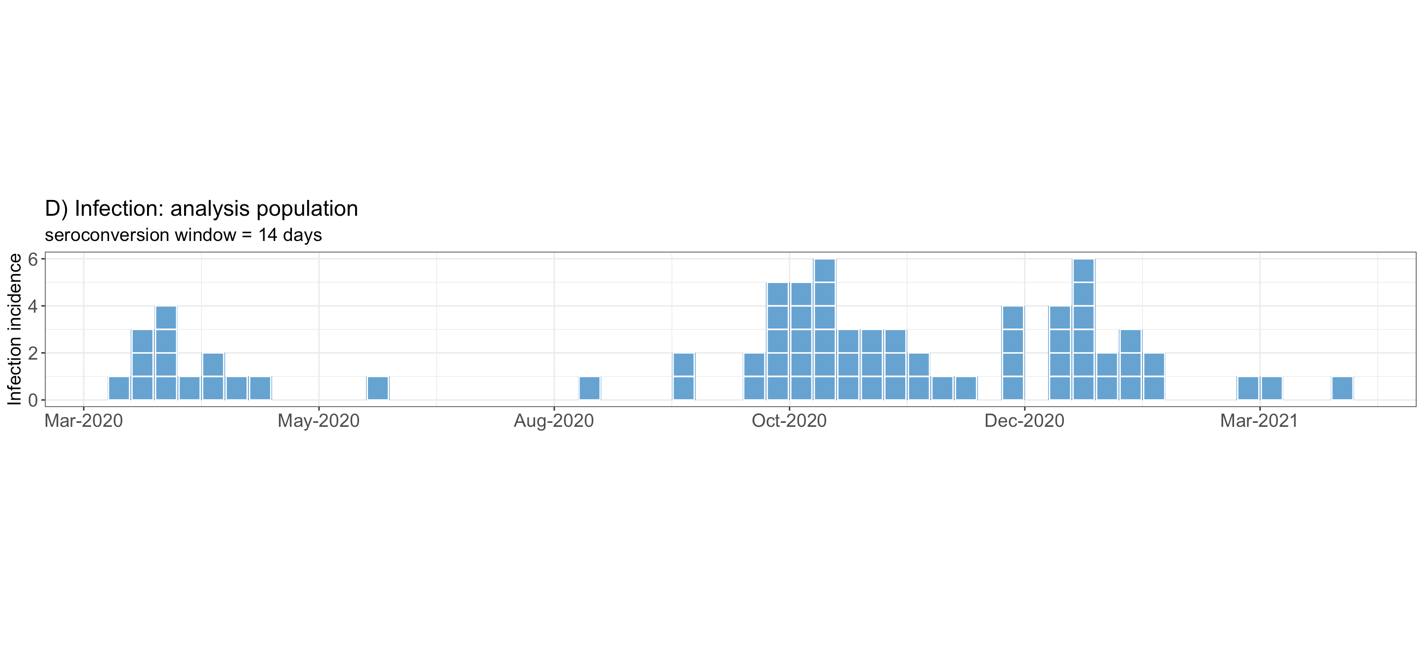

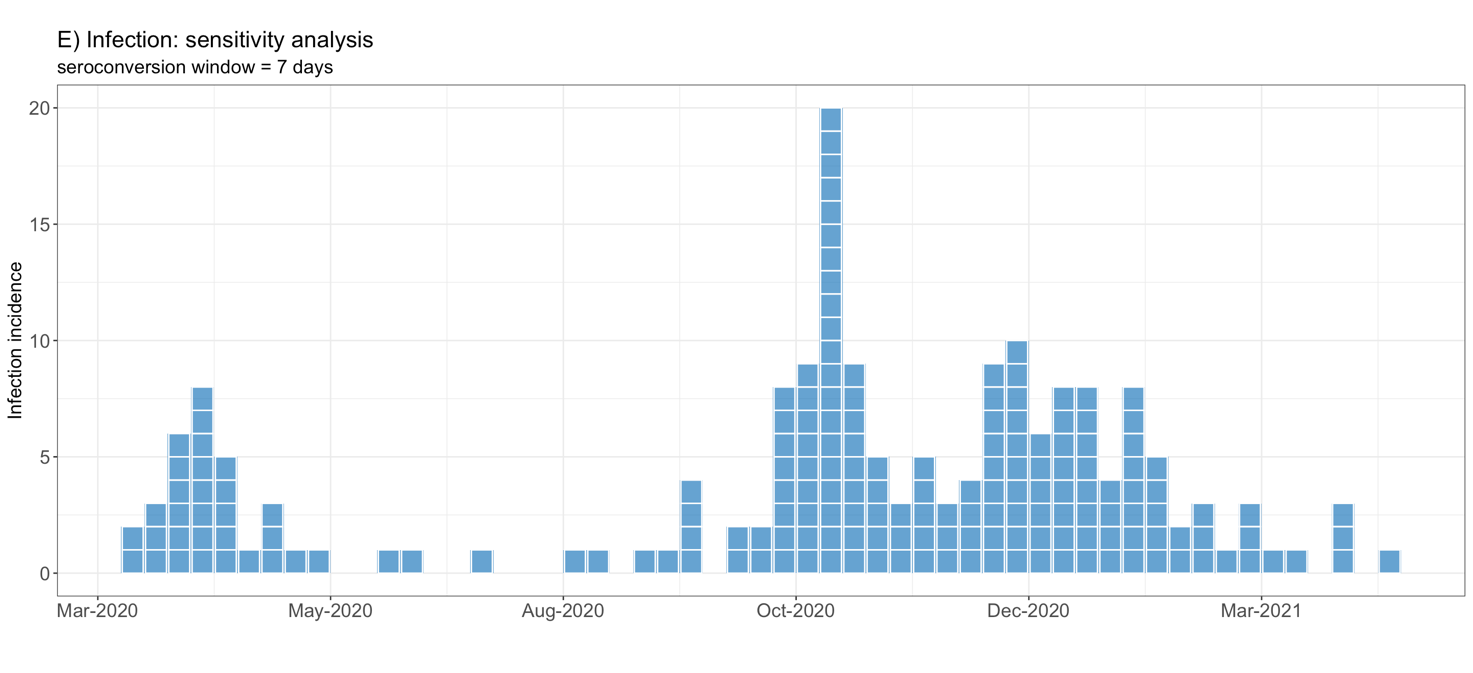

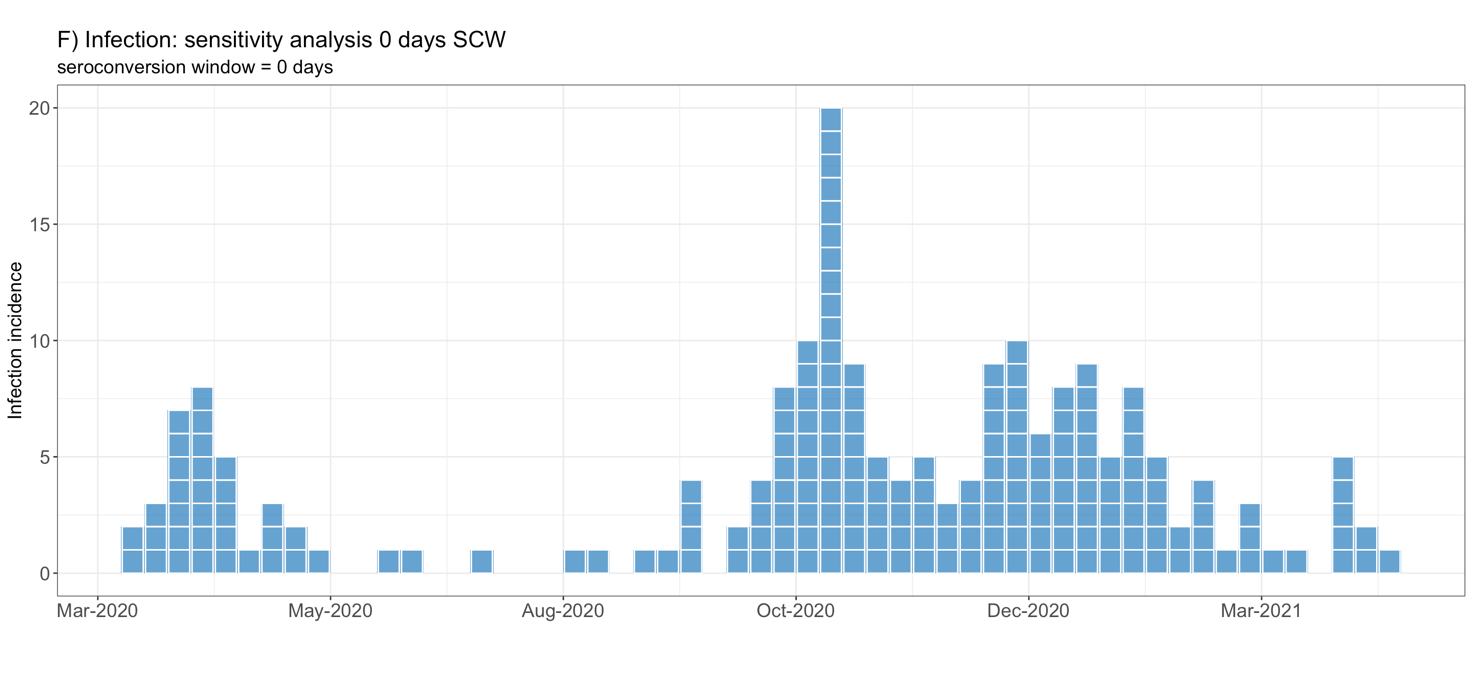

**A+D.** Window period is 14 days. The analysis population includes 970 participants and 223 infections. Four participants were in the seroconversion window for an infection and 125 for the first dose of a vaccine.

**B+E.** Window period is 7 days. The sensitivity analysis population includes 1,024 participants and 236 infections. Four participants were in the seroconversion window for an infection and 71 for the first dose of a vaccine.

**C+F.** Window period is 0 days. The sensitivity analysis population includes 1,099 participants and 249 infections.

**Table S1:	Participant baseline characteristics and immune events during follow-up with participants in 14-day seroconversion window removed from both populations**

| **A. Baseline characteristics**  Cells contain n (% of N) unless stated otherwise | **Analysis population**  **N= 970** | **Randomised population N=1,370^1^** | **p^2,3^** |
| --- | --- | --- | --- |
| **Age in years, mean (SD)** | 42.51 (12.59) | 41.57 (12.63) | 0.076 |
| **Female** | 724 (74.7) | 1003 (73.2) | 0.468 |
| **Work-related exposure^4^** Low  Medium  High | 148 (15.3)  240 (24.7)  582 (60.0) | 196 (62.3)  321 (23.4)  853 (62.3) | 0.540 |
| **Smoking status** Current  Former  Never | 65 (6.7)  302 (31.1)  603 (62.2) | 113 (8.2)  393 (28.7)  864 (63.1) | 0.222 |
| **History of BCG vaccination** | 174 (17.9) | 234 (17.1) | 0.629 |
| **Past tuberculosis test results^5^** Negative  Positive (either or both)  Never tested  Unknown (both) | 640 (66.0)  94 (9.7)  224 (23.1)  12 (1.2) | 922 (67.3)  124 (9.1)  312 (22.8)  12 (0.9) | 0.766 |
| **Respiratory infection in winter 2019-2020**  No  Yes, with fever  Yes, no fever | 713 (73.5)  79 (8.1)  178 (18.3) | 1000 (73.0)  115 (8.4)  255 (18.6) | 0.959 |
| **Influenza vaccination in winter 2020-2021^6^**  Yes  No  Missing | 528 (54.4)  311 (32.1)  131 (13.5) | 613 (44.7)  388 (28.3)  369 (26.9) | **<0.001** |
| **Influenza vaccination prior to follow-up** | 560 (57.7) | 798 (58.2) | 0.836 |
| **Any other vaccination in past year^7^** | 105 (10.8) | 154 (11.2) | 0.803 |
| **Current use of anti-hypertensive medication** | 61 (6.3) | 87 (6.4) | 1.000 |
| **History of cardiovascular disease** | 23 (2.4) | 32 (2.3) | 1.000 |
| **Current use of anti-diabetic medication** | 5 (0.5) | 7 (0.5) | 1.000 |
| **History of asthma** | 69 (7.1) | 95 (6.9) | 0.932 |
| **History of hay fever** | 288 (29.7) | 410 (29.9) | 0.938 |
| **History of other pulmonary disease** | 23 (2.4) | 30 (2.2) | 0.881 |
| **Any lung disease (previous three combined)** | 329 (33.9) | 462 (33.6) | 0.957 |
| **B. Immune events**  Cells contain n (% of N) unless stated otherwise | **Analysis population**  **N= 970** | **Randomised population**  **N=1,370^1^** | **p^2,3^** |
| **# Immune events^8^** 0  1  2  3 | 453 (46.7)  187 (19.3)  260 (26.8)  70 (7.2) | 679 (50.7)  223 (16.7)  351 (26.2)  86 (6.4) | 0.202 |
| **Immune event type** None  Infection^9^  Vaccine 1 dose  Vaccine 2 doses  Infection + 1 dose  Infection + 2 doses | 453 (46.7)  123 (12.7)  64 (6.6)  230 (23.7)  30 (3.1)  70 (7.2) | 679 (49.6)  152 (11.1)  57 (5.2)  320 (23.4)  30 (2.2)  86 (6.3) | **<0.001** |
| **COVID-19 vaccine product/dose^10^** None  mRNA 1 dose  mRNA 2 doses  Vector 1 dose  Vector 2 doses  Unknown 1 dose | 576 (59.4)  55 (5.7)  300 (30.9)  36 (3.7)  0 (0.0)  3 (0.3) | 862 (62.9)  58 (4.2)  399 (29.1)  38 (2.8)  7 (0.5)  6 (0.4) | 0.052 |
| **Had SARS-CoV-2 infection during follow-up^11^** | 223 (21.2) | 268 (20.3) | 0.094 |
| **Infection severity^12^** No infection  Asymptomatic  Very Mild  Mild  Moderate  Unknown | 747 (77.0)  33 (3.4)  120 (12.4)  44 (4.5)  2 (0.2)  24 (2.5) | 1102 (80.4)  38 (2.8)  148 (10.8)  52 (3.8)  2 (0.1)  28 (2.0) | 0.527 |
| **Acute duration of infection^13^** 0 days/no infection  0-1 weeks  1- 2 weeks  2-3 weeks  3-4 weeks  4+ weeks  Lingering  Ongoing/unknown | 782 (80.6)  29 (3.0)  45 (4.6)  32 (3.3)  17 (1.8)  15 (1.5)  22 (2.3)  28 (2.9) | 1136 (82.9)  40 (2.9)  52 (3.8)  41 (3.0)  18 (1.3)  16 (1.2)  28 (2.0)  39 (2.8) | 0.885 |
| **Long COVID** No  Yes  Unknown | 193 (86.5)  19 (8.5)  11 (4.9) | 230 (85.8)  23 (8.6)  15 (5.6) | 0.947 |
| **Long term loss of smell/taste** No  Yes  Unknown | 211 (94.6)  6 (2.7)  6 (2.7) | 252 (94.0)  8 (3.0)  8 (3.0) | 0.962 |

Abbreviations: BCG= Bacillus Calmette-Guerin; M=Month; SD=standard deviation.

1. Participants that were in the 14-day seroconversion window for an infection (N=4) or the first dose of a COVID-19 vaccination (N=133) were removed from the randomised population. An additional 17 participants were in the 14-day seroconversion window for a second COVID-19 vaccination. They were recoded as having received only one dose.
2. Chi-squared tests for categorical variables and Wilcoxon rank sum test for continuous variables.
3. Statistical test comparing baseline characteristics between the analysis (N=970) and randomised populations (N=1,370).
4. Work related exposure is a combination of participants expected to work in a COVID-19 ward and the percentage of hours with direct patient contact (Supplementary Methods).
5. Tuberculosis tests include the Mantoux and/or TB QuantiFERON tests.
6. Only the missing category is statistically significantly different between the analysis and randomised populations.
7. Included DTaP-IPV, hepatitis A, hepatitis B, yellow fever, typhoid, rabies, mumps-measles-rubella, meningococcal, pneumococcal, *Haemophilus influenza* type B, Ebola, tick-borne encephalitis, human papillomavirus, and unknown.
8. An immune event is considered to be either one SARS-CoV-2 infection or one dose of a COVID-19 vaccine.
9. No-one in the analysis population, and four persons in the randomised population, had more than one infection (two each).
10. COVID-19 vaccines available in the Netherlands during the study period are listed in the methods. In addition, one person received an experimental mRNA vaccine by CureVac N.V. in a clinical trial setting. That vaccine was never marketed due to insufficient efficacy, but the person was included in the mRNA vaccines group.
11. A natural infection was defined as a reported positive test (by the participants through the diary app) or identified by serology (evidence of anti-S1 at sampling round 1 or both anti-S1 and anti-N at sampling round 2). None of the participants in the analysis population reported a positive PCR test at baseline.
12. Participants with an unsure infection status at M12 (never reported a positive test and/or seropositive for anti-N but not anti-S1 at M12; n=36) were considered as never having had an infection during follow-up in the randomised population and were removed from the analysis population.
13. Participants who never had an infection were included in the 0 days/no infection category.

**Figure S3:	M12 anti-S1 and anti-N log_10_ concentrations for BCG versus placebo stratified by number of immune events**

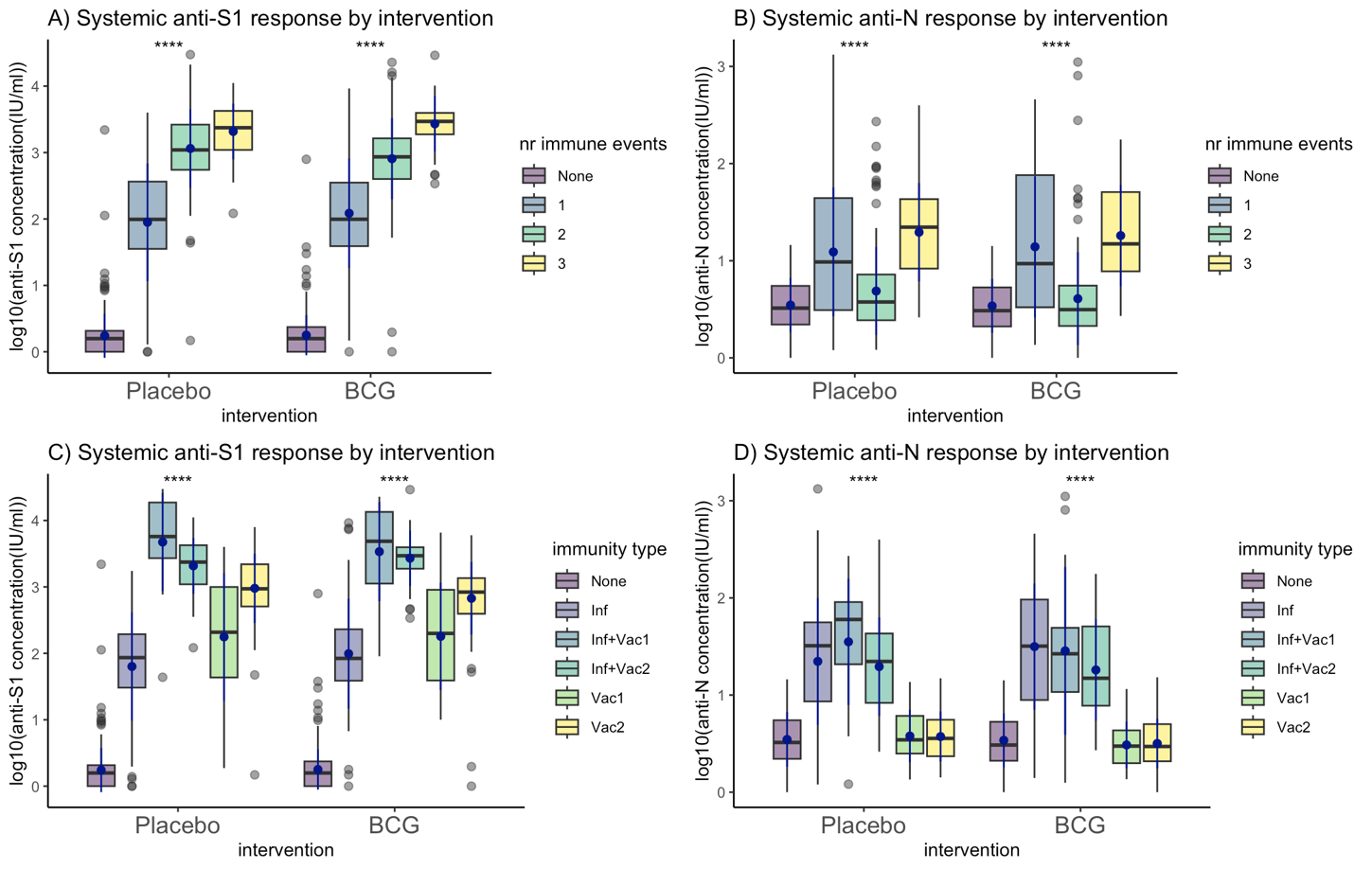

Abbreviations: Inf=infection; Inf+Vac1= infection and one dose of a COVID vaccine; Inf+Vac2= infection and two doses of a COVID vaccine; Vac1=one dose of a COVID vaccine; Vac2=two doses of a COVID vaccine.

Systemic antibody concentrations were log_10_ transformed (after adding a pseudocount of 1). An immune event was defined as one SARS-CoV-2 infection or one dose of a COVID-19 vaccine. The blue point range indicates the mean and standard deviation of the log_10_ concentrations. Statistical testing (****) shows the significance for within group comparisons. All statistical testing between BCG and placebo groups were non-significant.

**Figure S4:	Spearman correlations between M12 anti-S1 and anti-N log_10_ concentrations and time since SARS-CoV-2 infection (A-B) or time since first dose of a COVID-19 vaccine (C)**

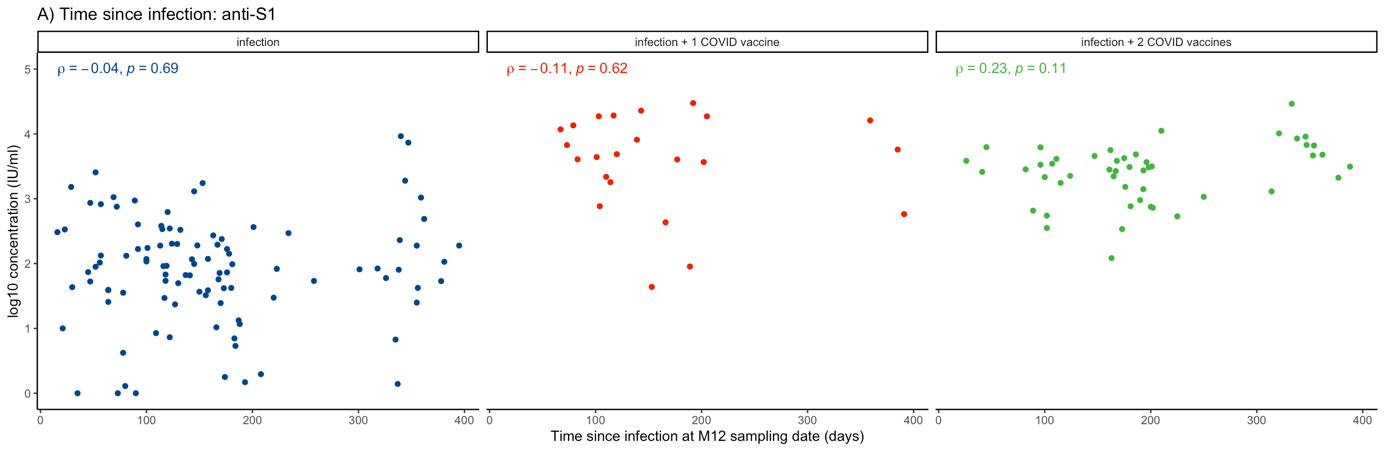

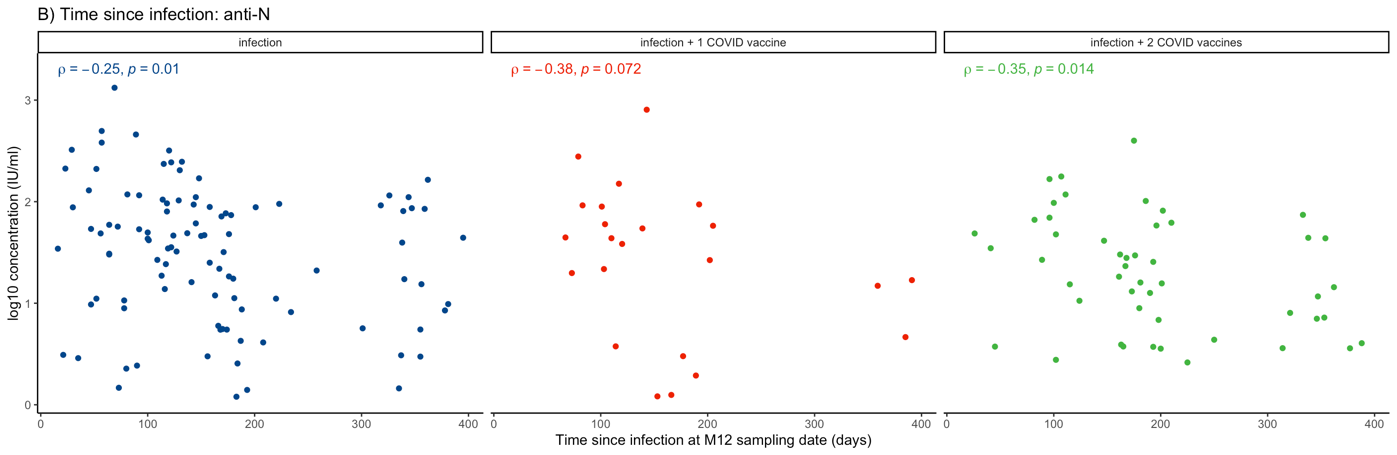

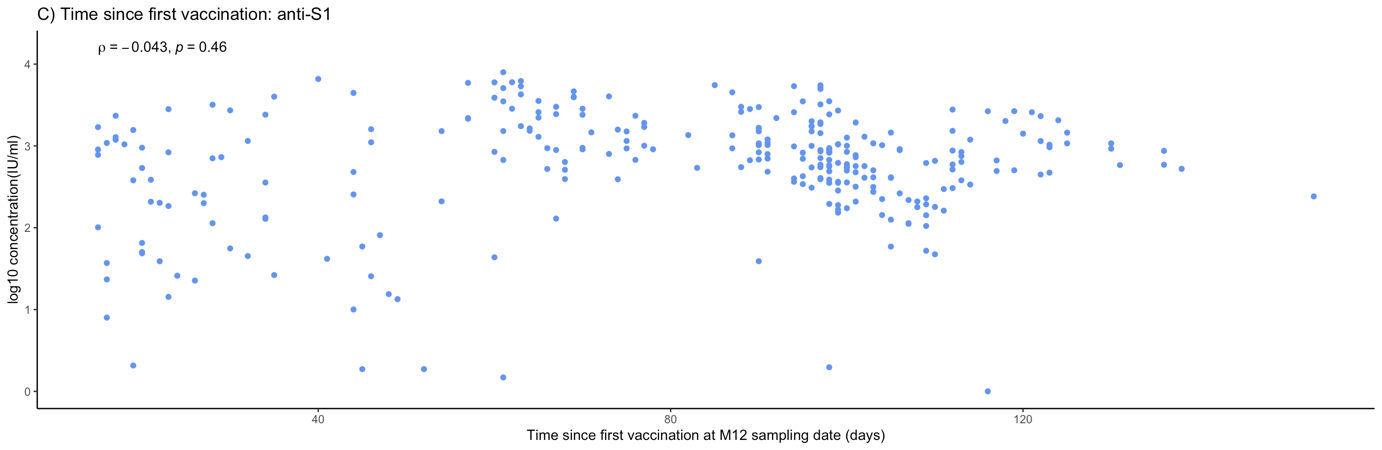

**A-B.** N=174; 49 participants with an unknown infection date excluded.

**C**. N=294; only includes participants who received at least one dose of a COVID-19 vaccine but never experienced a SARS-CoV-2 infection.

**Table S2: Mean M12 anti-S1 log_10_ concentrations by COVID-19 vaccine dose and type**

|  | | **M12 anti-S1** | | **M12 anti-N** | |
| --- | --- | --- | --- | --- | --- |
| **Vaccine type and dose^1^** | **N=747^2^ (%)** | **Mean log_10_ concentration**  **(SD) in IU/ml** | **Geometric mean (SD)^3^**  **in IU/ml** | **Mean log_10_ concentration (SD) in IU/ml** | **Geometric mean (SD)^3^**  **in IU/ml** |
| **None** | 453 (60.6) | 0.25 (0.32) | 1.78 (2.09) | 0.54 (0.28) | 3.47 (1.91) |
| **Comirnaty 1** | 4 (0.54) | 2.08 (0.66) | 120.23 (4.57) | 0.44 (0.10) | 2.75 (1.26) |
| **Comirnaty 2** | 226 (30.3) | 2.90 (0.54) | 794.33 (3.47) | 0.53 (0.26) | 3.39 (1.82) |
| **Spikevax 1** | 31 (4.1) | 2.75 (0.75) | 562.34 (5.62) | 0.57 (0.29) | 3.72 (1.95) |
| **Spikevax 2** | 3 (0.40) | 3.09 (0.32) | 1230.23 (2.09) | 0.63 (0.29) | 4.27 (1.95) |
| **Vaxzevria 1** | 25 (3.4) | 1.78 (0.73) | 60.26 (5.37) | 0.51 (0.23) | 3.24 (1.70) |
| **Jcovden 1** | 3 (0.40) | 1.93 (0.25) | 85.11 (1.78) | 0.55 (0.34) | 3.55 (2.19) |
| **CureVac 2** | 1 (0.13) | 2.11 (NA) | 128.82 (NA) | 0.65 (NA) | 4.47 (NA) |
| **Unknown 1** | 1 (0.13) | 0.32 (NA) | 2.09 (NA) | 0.24 (NA) | 1.74 (NA) |

Abbreviations: IU/ml= international unit per millilitre; M=month, SD=standard deviation.

1. In the Netherlands, the COVID-19 vaccines available during the study period were Spikevax (Moderna Biotech, Cambridge, MA, USA), Comirnaty (Pfizer/BioNTech, New York, NY, USA), Vaxzevria (AstraZeneca AB, Sodertalje, Sweden), and Jcovden (Janssen Vaccines, Leiden, Netherlands). In addition, one participant received an experimental vaccine by CureVac N.V. in a clinical trial setting. This vaccine was never marketed due to insufficient efficacy.
2. N=747; participants who experienced an infection only were excluded (n=223).
3. Geometric means are the antilog 10^x^ of the log_10_ concentrations.

**Figure S5:	Mean M12 anti-S1 and anti-N log_10_ concentrations for infection episode duration and individual symptom severity and duration**

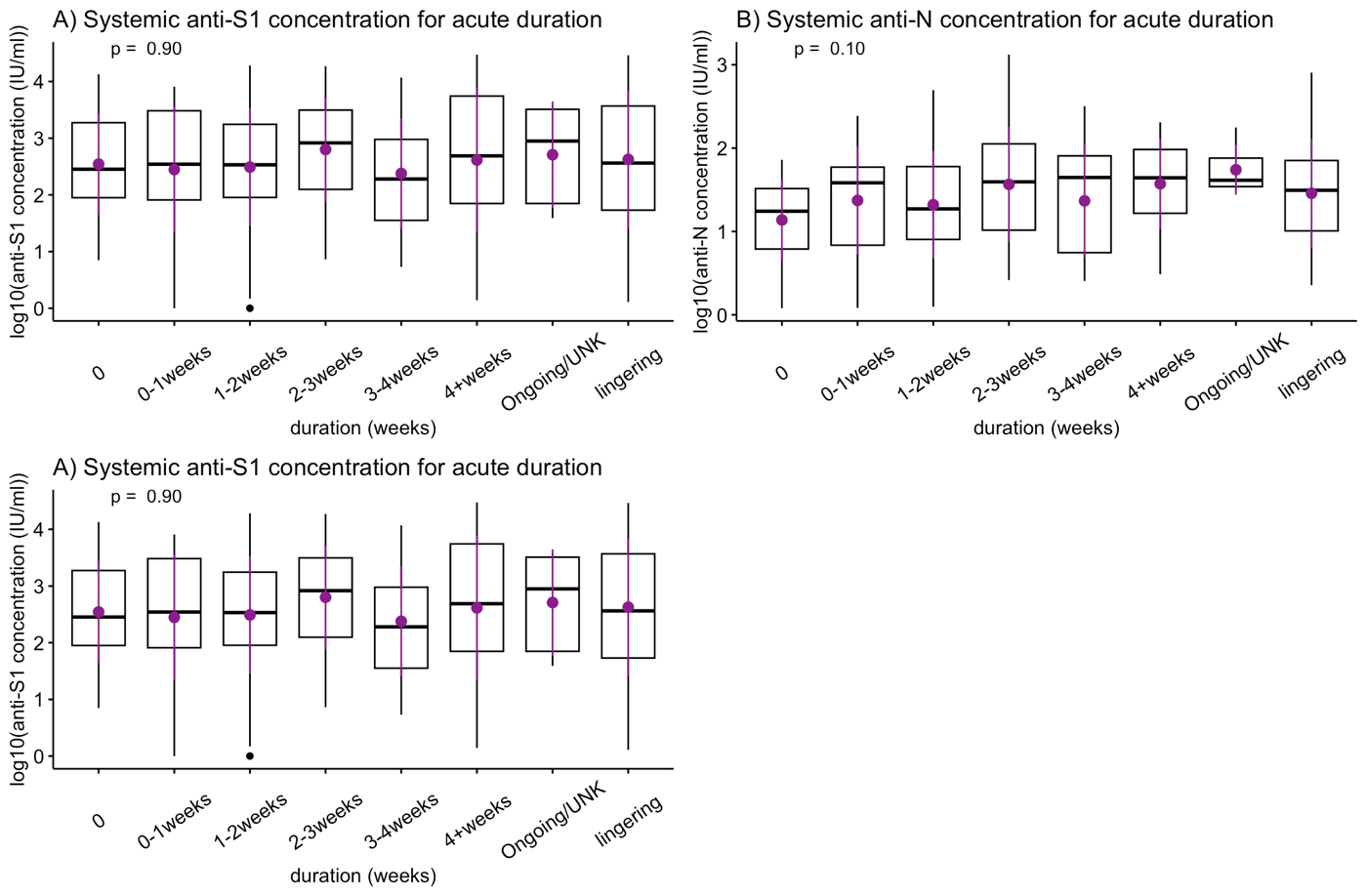

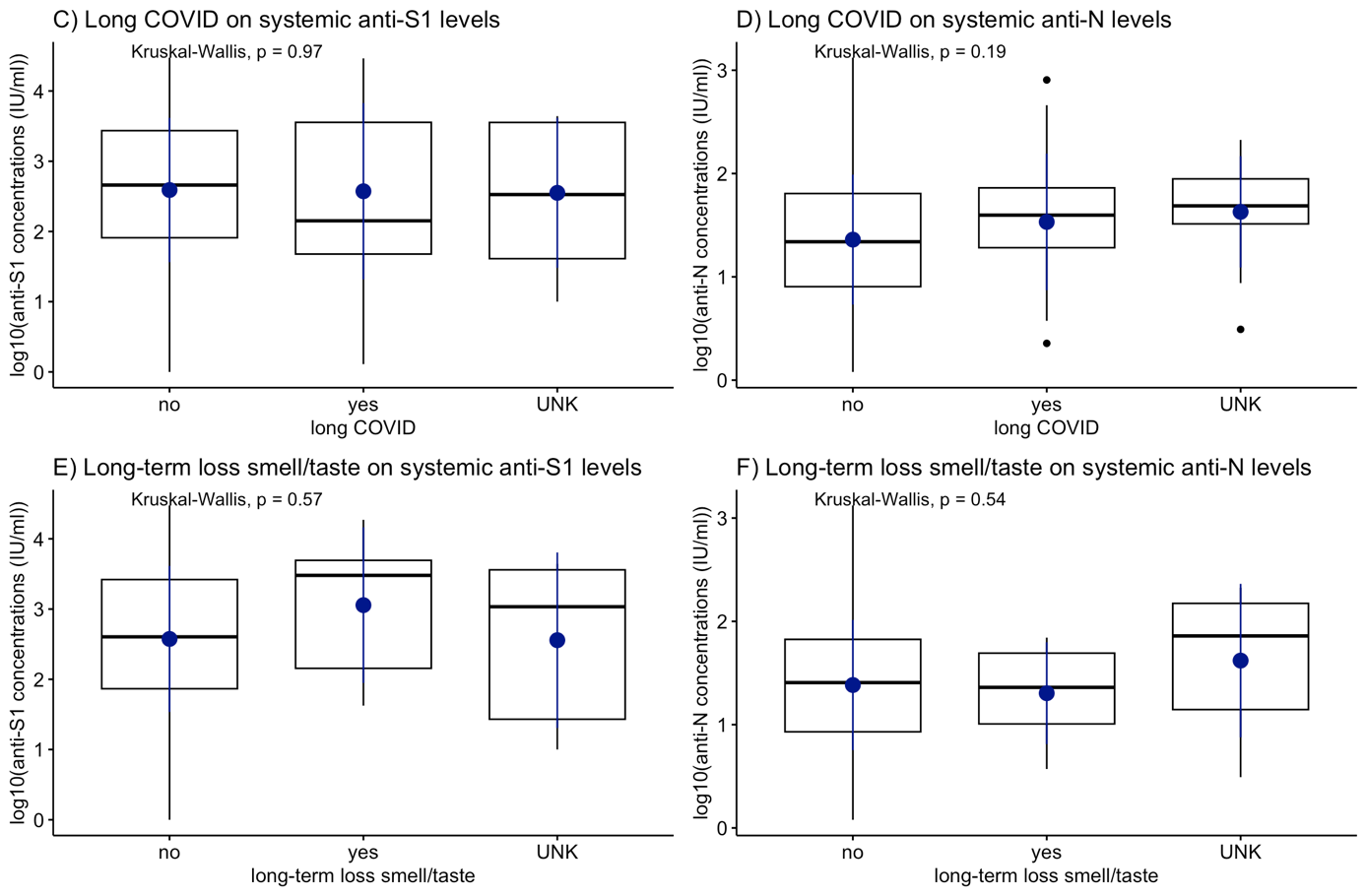

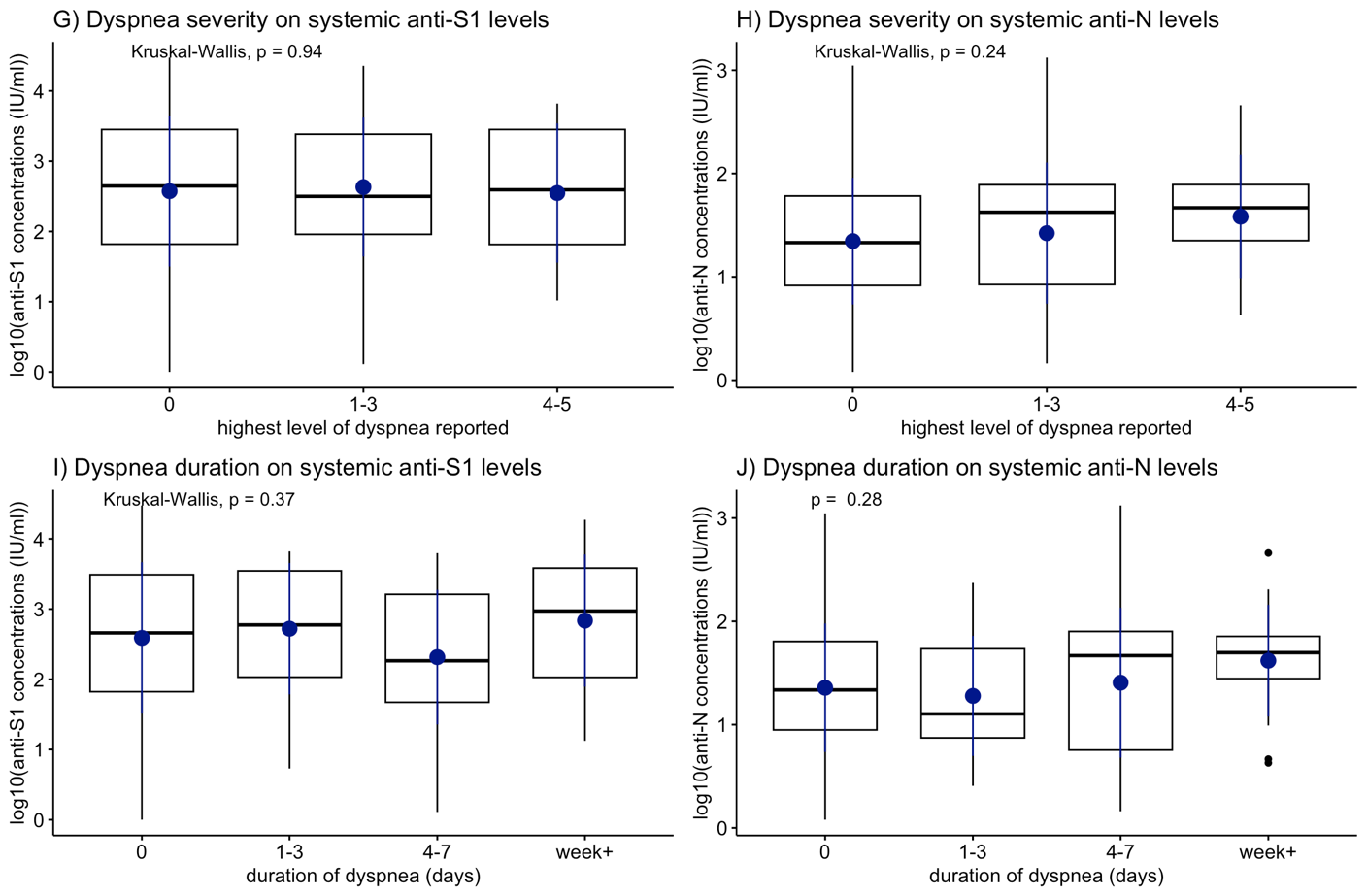

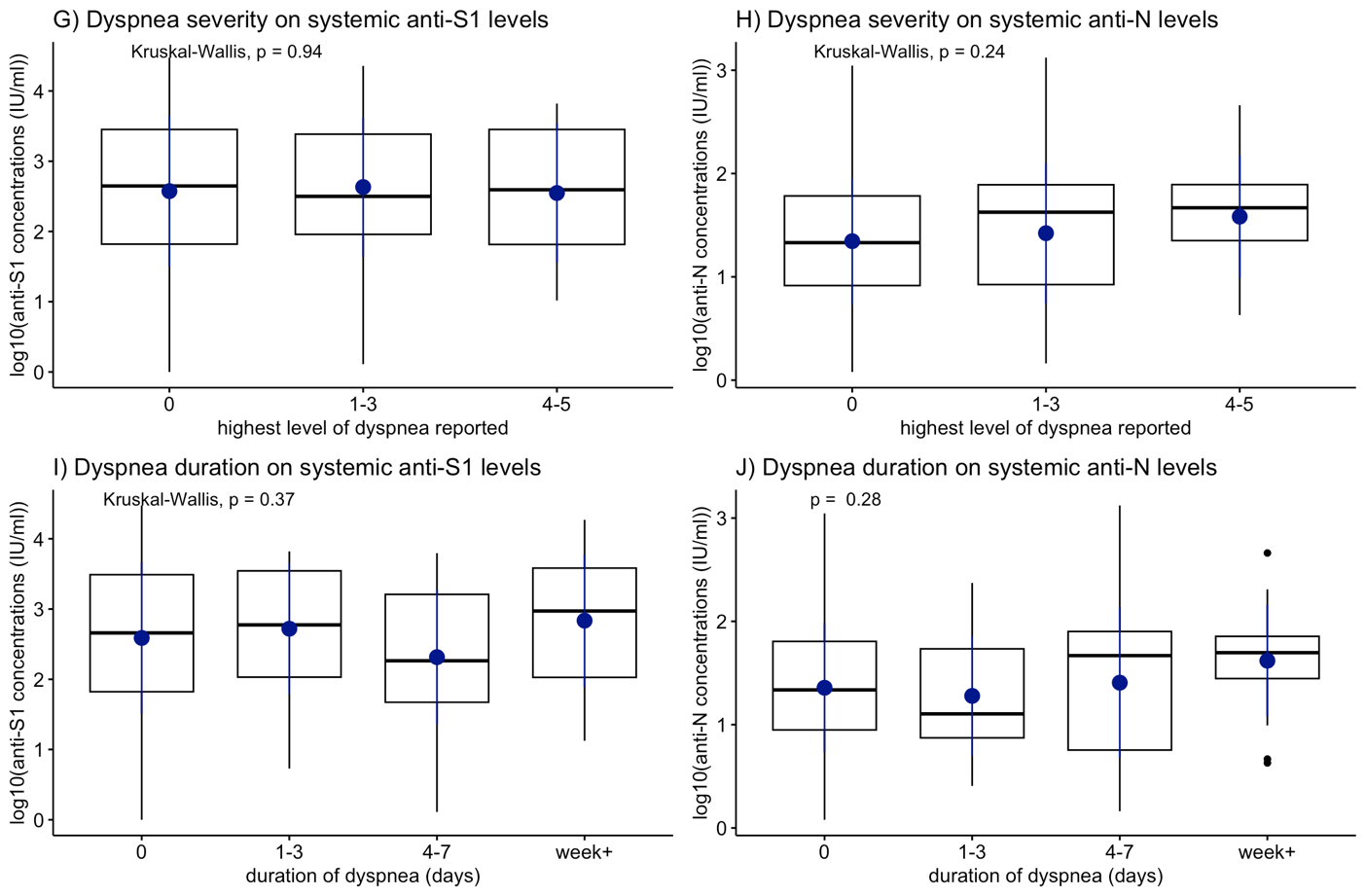

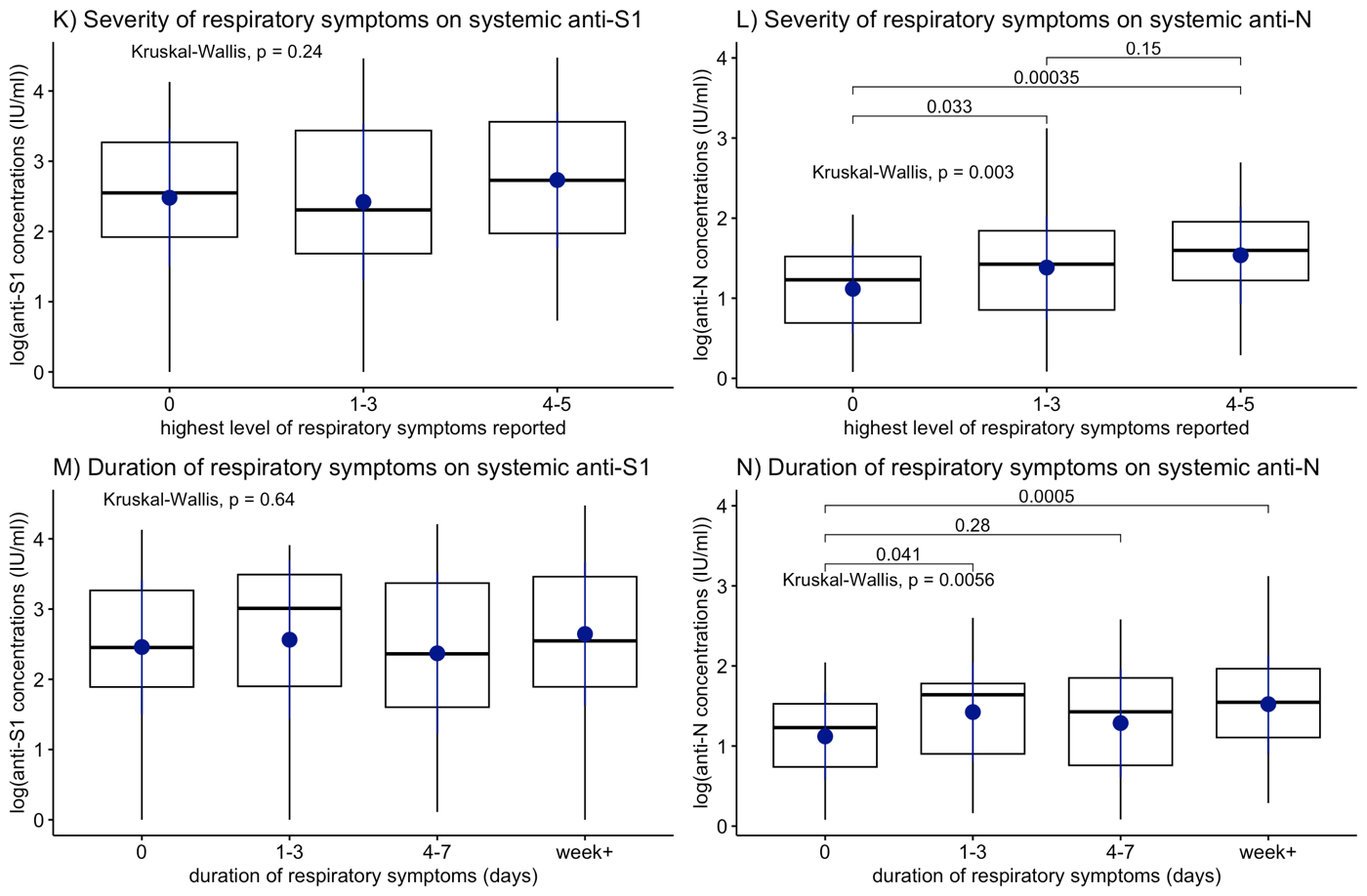

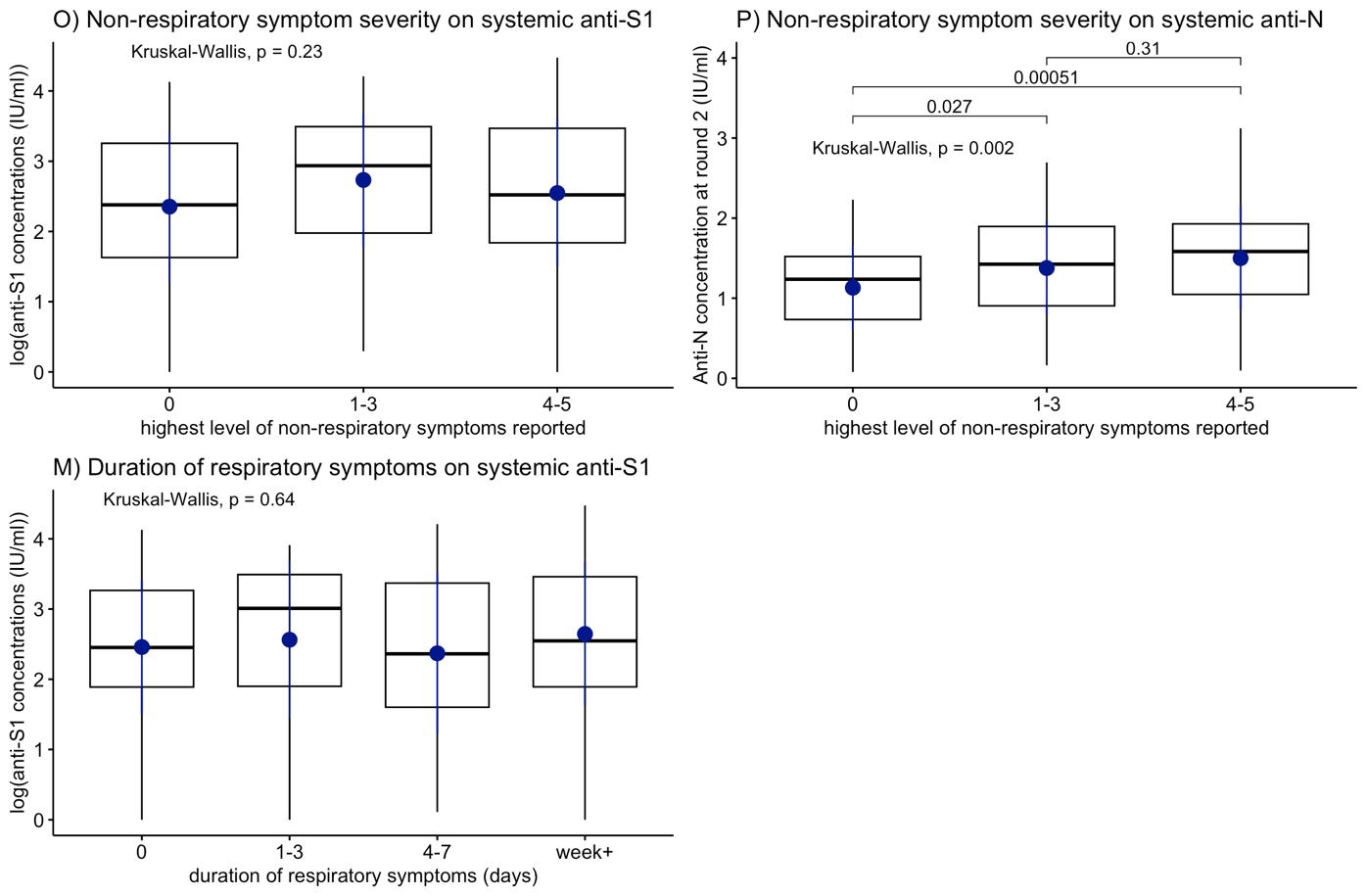

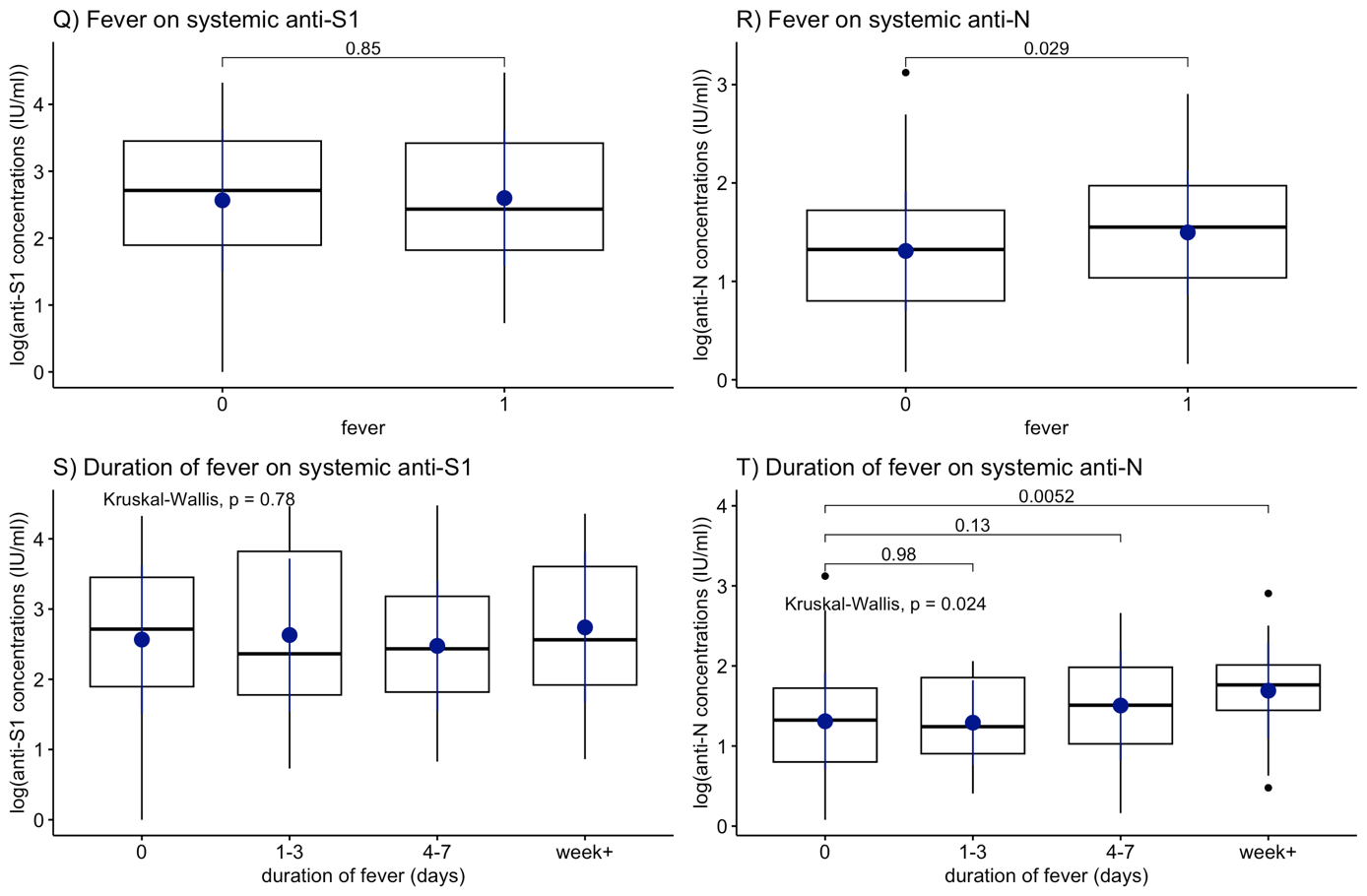

Systemic antibody concentrations were log_10_ transformed (after adding a pseudocount of 1).

**A-B.** Acute duration is defined as the days between the start of symptoms and end of symptoms surrounding an infection, excluding lingering non-respiratory symptoms. All infections that had an uncertain date (because the infection was identified through serology) and all ongoing infections at the end of follow-up were classified into the ongoing/unknown group. All infections with lingering symptoms, such as Long COVID or long-term loss of smell/taste, were classified as lingering.

**C-F.** Long COVID was defined as continuing to report symptoms other than standalone loss of smell/taste for at least 60 days after the end of the acute infection episode. Long-term loss of taste/smell was defined separately from Long COVID, as continuing to report standalone loss of smell/taste for at least 60 days after the end of the acute infection episode. Participants who were actively reporting symptoms past the end of their acute episode duration when the app was terminated, but had not yet reached two months were coded as unknown.

**G-P**. Severity was reported on a scale of 0-5, 0 indicating not present and 1-5 in increasing severity. Duration of symptoms is the number of days that the particular symptom was reported within the acute episode duration.

**Q-T.** Fever was defined as temperatures above 38°C.

**Figure S6:	Spearman correlations between M12 anti-S1 and anti-N log_10_ concentrations in participants who had a SARS-CoV-2 infection plus no, one or two vaccinations**

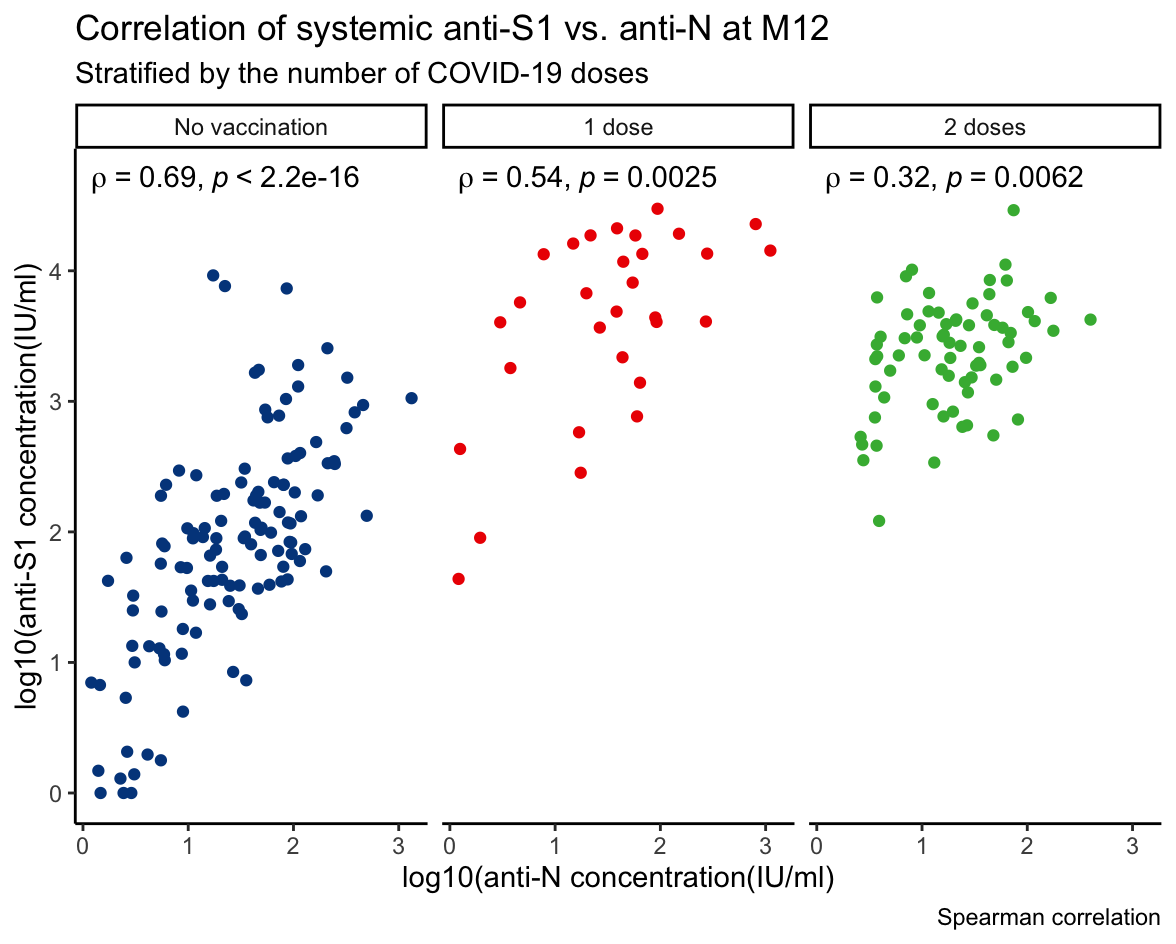

Systemic antibody concentrations were log_10_ transformed (after adding a pseudocount of 1).

**Table S3: Univariable linear regression results for the full analysis population (A) and participants who experienced an infection (B)**

| **A. Analysis population^1^** | **M12 anti-S1 log_10_ concentration** | | | | **M12 anti-N log_10_ concentration** | | | |
| --- | --- | --- | --- | --- | --- | --- | --- | --- |
|  | **Intercept**  **(95% CI)** | **Model estimate**  **(95% CI)** | **p** | **R^2^** | **Intercept**  **(95% CI)** | **Model estimate**  **(95% CI)** | **p** | **R^2^** |
| **Participant characteristics** | | | | | | | | |
| Age (in years) | 1.73 (1.43, 2.04) | -0.004 (-0.01, 0.003) | 0.216 | 0.00 | 0.74 (0.63, 0.86) | -0.002 (-0.003, 0.002) | 0.861 | 0.00 |
| Male sex | 1.52 (1.42, 1.62) | 0.11 (-0.09, 0.31) | 0.271 | 0.00 | 0.73 (0.69, 0.77) | 0.02 (-0.06, 0.09) | 0.625 | 0.00 |
| Smoking Never  Current  Former | 1.56 (1.45, 1.67) | -0.04 (-0.40, 0.31)  -0.03 (-0.22, 0.16) | 0.816  0.771 | 0.00 | 0.73 (0.69, 0.78) | -0.07 (-0.21, 0.06)  0.01 (-0.05, 0.09) | 0.274  0.722 | 0.00 |
| Work-related exposure^2^ Low  Medium  High | 0.67 (0.46, 0.87) | 0.39 (0.13,0.65)  1.30 (1.07, 1.53) | **0.004**  **<0.001** | 0.15 | 0.67 (0.58, 0.75) | 0.03 (-0.07, 0.14)  0.10 (0.00, 0.10) | 0.551  **0.049** | 0.01 |
| Additional household members | 1.55 (1.40, 1.70) | -0.002 (-0.07, 0.06) | 0.956 | 0.00 | 0.72 (0.66, 0.77) | 0.01 (-0.01, 0.03) | 0.453 | 0.00 |
| History of BCG vaccination | 1.54 (1.44, 1.63) | 0.06 (-0.17, 0.28) | 0.623 | 0.00 | 0.72 (0.68, 0.76) | 0.08 (-0.01, 0.16) | 0.082 | 0.00 |
| Past TB test results^3^ Negative  Positive  Not done  Unknown | 1.60 (1.50, 1.71) | 0.12 (-0.18, 0.42)  -0.25 (-0.46, -0.04)  -0.86 (-1.65, -0.07) | 0.447  **0.020**  **0.033** | 0.01 | 0.72 (0.68, 0.76) | 0.06 (-0.05, 0.18)  0.02 (-0.06, 0.10)  0.11 (-0.19, 0.41) | 0.284  0.557  0.466 | 0.00 |
| Respiratory infection in winter 2019-2020^4^  No  Yes, with fever  Yes, no fever | 1.55 (1.45, 1.66) | -0.17 (-0.49, 0.15)  0.03 (-0.20, 0.26) | 0.299  0.785 | 0.00 | 0.74 (0.70, 0.78) | -0.02 (-0.14, 0.10)  -0.04 (-0.13, 0.04) | 0.719  0.344 | 0.00 |
| Influenza vaccine in winter 2019-2020  No  Yes  Missing | 1.57 (1.41, 1.72) | -0.13 (-0.32, 0.07)  0.37 (0.09, 0.65) | 0.196  **0.010** | 0.01 | 0.75 (0.69, 0.81) | -0.04; (-0.12, 0.03)  0.03; (-0.08, 0.14) | 0.242  0.594 | 0.00 |
| Influenza vaccine prior to follow-up | 1.61 (1.47, 1.74) | -0.10 (-0.28, 0.08) | 0.260 | 0.00 | 0.73 (0.68, 0.78) | -0.002 (-0.07, 0.07) | 0.995 | 0.00 |
| Any other vaccination in past year^5^ | 1.56 (1.47, 1.66) | -0.16 (-0.45, 0.12) | 0.251 | 0.00 | 0.74 (0.70, 0.77) | -0.05 (-0.16, 0.05) | 0.333 | 0.00 |
| Current use of anti-hypertensive medication | 1.55 (1.46, 1.64) | -0.05 (-0.41, 0.31) | 0.767 | 0.00 | 0.72 (0.69, 0.75) | 0.20 (0.07, 0.39) | **0.003** | 0.00 |
| Current use of anti-diabetic medication | 1.55 (1.46, 1.63) | 0.10 (-1.12, 1.32) | 0.871 | 0.00 | 0.73 (0.70, 0.77) | -0.27 (-0.73, 0.19) | 0.249 | 0.00 |
| History of cardiovascular disease | 1.54 (1.45, 1.63) | 0.22 (-0.36, 0.79) | 0.463 | 0.00 | 0.73 (0.69, 0.76) | 0.19 (-0.02, 0.41) | 0.079 | 0.00 |
| History of asthma | 1.54 (1.45, 1.63) | 0.09 (-0.25, 0.43) | 0.585 | 0.00 | 0.73 (0.70, 0.77) | -0.01 (-0.14, 0.12) | 0.912 | 0.00 |
| History of hay fever | 1.57 (1.47, 1.68) | -0.09 (-0.28, 0.10) | 0.336 | 0.00 | 0.73 (0.69, 0.77) | 0.01 (-0.06, 0.09) | 0.701 | 0.00 |
| History of other pulmonary disease | 1.54 (1.45, 1.63) | 0.32 (-0.25, 0.90) | 0.273 | 0.00 | 0.73 (0.69, 0.76) | 0.24 (0.02, 0.46) | **0.031** | 0.00 |
| Any lung disease (previous three combined) | 1.56 (1.45, 1.67) | -0.04 (-0.23, 0.14) | 0.666 | 0.00 | 0.73 (0.69, 0.77) | 0.02 (-0.05, 0.09) | 0.667 | 0.00 |
| Intervention: BCG | 1.59 (1.46, 1.71) | -0.08 (-0.26, 0.09) | 0.355 | 0.00 | 0.75 (0.71, 0.80) | -0.04 (-0.11, 0.03) | 0.235 | 0.00 |
| **Immune events** | | | | | | | | |
| Had a SARS-CoV-2 infection during follow-up | 1.24 (1.14, 1.33) | 1.35 (1.16, 1.54) | **<0.001** | 0.17 | 0.54 (0.51, 0.56) | 0.85 (0.79, 0.91) | **<0.001** | 0.47 |
| Acute infection duration^6^ 0 days/no infection  0-1 weeks  1- 2 weeks  2-3 weeks  3-4 weeks  4+ weeks  Lingering  Ongoing/unknown | 1.29 (1.20, 1.38) | 1.15 (0.67, 1.63)  1.20 (0.81, 1.58)  1.51 (1.05, 1.96)  1.08 (0.46, 1.71)  1.32 (0.66, 1.98)  1.33 (0.79, 1.88)  1.50 (1.02, 1.99) | **<0.001**  **<0.001**  **<0.001**  **0.001**  **<0.001**  **<0.001**  **<0.001** | 0.14 | 0.56 (0.54, 0.59) | 0.81 (0.66, 0.95)  0.76 (0.64, 0.88)  1.00 (0.86, 1.14)  0.80 (0.61, 0.99)  1.01 (0.81, 1.21)  0.89 (0.73, 1.06)  0.93 (0.78, 1.08) | **<0.001**  **<0.001**  **<0.001**  **<0.001**  **<0.001**  **<0.001**  **<0.001** | 0.44 |
| Number of immune events^7^ 0  1  2  3 | 0.25; (0.20, 0.30) | 1.77 (1.68, 1.86)  2.74 (2.65, 2.82)  3.12 (2.98, 3.25) | **<0.001**  **<0.001**  **<0.001** | 0.84 | 0.54; (0.50, 0.58) | 0.58 (0.50, 0.66)  0.11 (0.05, 0.18)  0.74 (0.63.0.86) | **<0.001**  **0.002**  **<0.001** | 0.25 |
| Immune event type None  Infection  Vaccine 1 dose  Vaccine 2 doses  Infection+1 dose  Infection+2 doses | 0.25; (0.20, 0.30) | 1.65 (1.55, 1.76)  2.00 (1.87, 2.14)  2.66 (2.57, 2.74)  3.37 (3.16, 3.55)  3.12 (2.98, 3.25) | **<0.001**  **<0.001**  **<0.001**  **<0.001**  **<0.001** | 0.85 | 0.54; (0.50, 0.57) | 0.88 (0.81, 0.96)  -0.008 (-0.11, 0.09)  -0.002 (-0.06, 0.06)  0.96 (0.82, 1.10)  0.74 (0.64, 0.84) | **<0.001**  0.880  0.943  **<0.001**  **<0.001** | 0.47 |
| Vaccine product and dose^8^ None  mRNA 1 dose  mRNA 2 doses  Vector 1  Unknown 1 dose | 0.60; (0.54, 0.66) | 2.49 (2.28, 2.70)  2.41 (2.30, 2.52)  1.46 (1.20, 1.72)  2.14 (1.28, 3.01) | **<0.001**  **<0.001**  **<0.001**  **<0.001** | 0.70 | 0.72; (0.68, 0.77) | 0.15 (0.004, 0.29)  -0.02 (-0.09, 0.06)  0.03 (-0.14, 0.21)  0.44 (-0.15, 1.04) | **0.044**  0.639  0.728  0.140 | 0.00 |
| Infection severity No infection  Asymptomatic  Very Mild  Mild  Moderate  Unknown | 1.24 (1.14, 1.33) | 1.31 (0.87, 1.75)  1.28 (1.04, 1.53)  1.44 (1.05, 1.82)  2.13 (0.37, 3.89)  1.54 (1.03, 2.06) | **<0.001**  **<0.001**  **<0.001**  **0.018**  **<0.001** | 0.17 | 0.54; (0.51, 0.56) | 0.60 (0.47, 0.73)  0.82 (0.75, 0.89)  1.11 (0.99, 1.23)  1.13 (0.60, 1.65)  0.85 (0.70, 1.00) | **<0.001**  **<0.001**  **<0.001**  **<0.001**  **<0.001** | 0.49 |

Abbreviations: CI= confidence interval; M=month.

1. N=970. Linear regression with log_10_ transformed concentrations of systemic M12 anti-S1 and anti-N (after a pseudo-count of 1 was added) as outcome. Each covariate was individually considered in a univariable linear regression. For categorical variables, indicator variables were first created, as to not assume a linear relation between the categories.
2. Work related exposure is a combination of participants expected to work in a COVID ward and the percentage of hours with direct patient contact.
3. Tuberculosis tests include the Mantoux and/or TB QuantiFERON tests.
4. No respiratory tract infection is the reference.
5. The following other vaccinations were reported: DTaP-IPV, hepatitis A, hepatitis B, yellow fever, typhoid, rabies, mumps-measles-rubella, meningococcal, pneumococcal, *Haemophilus influenza* type B, Ebola, tick-borne encephalitis, human papillomavirus, and unknown.
6. Participants who never had a SARS-CoV-2 infection are included in the 0 days/no infection category.
7. An immune event is considered to be either one SARS-CoV-2 infection or one dose of a COVID-19 vaccine.
8. COVID-19 vaccines available in the Netherlands during the study period are listed in the methods. In addition, one person received an experimental mRNA vaccine by CureVac N.V. in a clinical trial setting. That vaccine was never marketed due to insufficient efficacy, but the person was included in the mRNA vaccines group. One mRNA vaccine dose was significantly associated with M12 anti-N log_10_ concentration, but this is due to the fact that a greater proportion of participants in that group experienced an infection (36.4%, compared 23.3% and 22.2% in the groups who received two mRNA vaccine doses or one vector vaccine dose, respectively).

| **B. Participants with a SARS-CoV-2 infection^1^** | **M12 anti-S1 log_10_ concentration** | | | | **M12 anti-N log_10_ concentration** | | | |
| --- | --- | --- | --- | --- | --- | --- | --- | --- |
|  | **Intercept**  **(95% CI)** | **Model estimate**  **(95% CI)** | **p** | **R^2^** | **Intercept**  **(95% CI)** | **Model estimate (95% CI)** | **p** | **R^2^** |
| Overall severity^2^ Asymptomatic  Very Mild  Mild  Moderate | 2.54 (2.19, 2.90) | -0.03 (-0.43, 0.38)  0.13 (-0.34, 0.60)  0.82 (-0.66, 2.31) | 0.898  0.587  0.277 | 0.00 | 1.14 (0.93, 1.35) | 0.22 (-0.02, 0.45)  0.51 (0.24, 0.79)  0.53 (-0.35, 1.40) | 0.070  **<0.001**  0.238 | 0.07 |
| Long COVID^3,4^ No  Yes  Ongoing/unknown | 2.59 (2.44, 2.74) | -0.02 (-0.51, 0.48)  -0.04 (-0.68, 0.60) | 0.943  0.901 | 0.00 | 1.36 (1.27, 1.45) | 0.17; (-0.11, 0.65)  0.27; (-0.13, 0.47) | 0.257  0.168 | 0.01 |
| Long term loss of smell/taste^3,5^ Yes  No  Ongoing/unknown | 2.57 (2.43, 2.72) | 0.48 (-0.87, 0.84)  -0.02 (-0.37, 1.33) | 0.268  0.967 | 0.01 | 1.38 (1.30, 1.47) | 0.24 (-0.28, 0.75)  -0.08 (-0.59, 0.44) | 0.762  0.366 | 0.00 |
| Acute infection duration^3,6^ 0 days  0-1weeks  1-2weeks  2-3weeks  3-4weeks  4+weeks  Lingering  Ongoing/unknown | 2.52 (2.18, 2.88) | -0.08 (-0.60, 0.44)  -0.04 (-0.50, 0.43)  0.27 (-0.23, 0.78)  -0.15 (-0.76, 0.46)  0.09 (-0.55, 0.73)  0.10 (-0.45, 0.66)  0.27 (-0.26, 0.79) | 0.758  0.873  0.290  0.629  0.791  0.730  0.314 | 0.02 | 1.13; (0.92, 1.34) | 0.24 (-0.07, 0.55)  0.19 (-0.08, 0.47)  0.44 (0.14, 0.74)  0.24 (-0.12, 0.60)  0.44 (0.06, 0.82)  0.33 (-0.01, 0.66)  0.36 (0.05, 0.67) | 0.122  0.172  **0.005**  0.197  **0.022**  0.054  **0.023** | 0.05 |
| Dyspnea severity^7,8^ 0  1-3  4-5 | 2.57 (2.41, 2.74) | 0.06 (-0.29, 0.41)  -0.03 (-0.57, 0.52) | 0.744  0.923 | 0.00 | 1.35 (1.25, 1.44) | 0.08 (-0.13, 0.29)  0.23 (-0.09, 0.56) | 0.470  0.150 | 0.01 |
| Duration of dyspnea^7^ (days, continuous) | 2.55 (2.41, 2.70) | 0.01 (-0.01, 0.03) | 0.233 | 0.01 | 1.36 (1.27, 1.44) | 0.01 (-0.002, 0.02) | 0.113 | 0.01 |
| Respiratory symptoms severity^9,10^ 0  1-3  4-5 | 2.48 (2.17, 2.79) | -0.07 (-0.44, 0.32)  0.25 (-0.15, 0.65) | 0.757  0.217 | 0.02 | 1.12; (0.93, 1.30) | 0.27 (0.04, 0.49)  0.42 (0.18, 0.66) | **0.019**  **0.001** | 0.06 |
| Respiratory symptoms duration^9,11^ (days) | 2.52 (2.32, 2.72) | 0.002 (-0.01, 0.02) | 0.743 | 0.00 | 1.26 (1.14, 1.38) | 0.01 (0.004, 0.02) | **0.005** | 0.04 |
| Fever (yes/no)^12^ | 2.57 (2.40, 2.74) | 0.03 (-0.26, 0.33) | 0.824 | 0.00 | 1.31 (1.21, 1.41) | 0.19 (0.01, 0.36) | **0.035** | 0.02 |
| Fever duration^12^ (days) | 2.57 (2.41, 2.73) | 0.003 (-0.03, 0.03) | 0.876 | 0.00 | 1.31 (1.22, 1.40) | 0.03 (0.01, 0.05) | **0.002** | 0.04 |
| Non-respiratory symptoms severity^13,14^ 0  1-3  4-5 | 2.35; (2.07, 2.64) | 0.38 (-0.03, 0.79)  0.19 (-0.15, 0.54) | 0.070  0.271 | 0.02 | 1.13 (0.96, 1.30) | 0.25 (0.003, 0.49)  0.37 (0.16, 0.58) | **0.048**  **<0.001** | 0.06 |

Abbreviations: CI=confidence interval; M=month.

1. Only those who experienced a SARS-CoV-2 infection were included. Linear regression with log_10_ transformed concentrations of systemic M12 anti-S1 and anti-N (after a pseudo-count of 1 was added) as outcome. Each covariate was individually considered in a univariable linear regression and categorical variables were considered ordinal.
2. N=199: Includes only participants who experienced a SARS-CoV-2 infection and an additional 24 participants were removed due to unknown infection severity. Includes only two moderate infections.
3. N=223: Includes only participants who experienced a SARS-CoV-2 infection.
4. Long COVID was defined as continuing to report symptoms other than standalone loss of smell/taste for at least 60 days after the end of the acute infection episode.
5. Long-term loss of taste/smell was defined as continuing to report standalone loss of smell/taste for at least 60 days after the end of the acute infection episode.
6. Acute duration of zero is the reference. Acute duration is defined as the days between the start of symptoms and end of symptoms surrounding an infection, excluding lingering non-respiratory symptoms. All infections that had an uncertain date (because the infection was identified through serology only) and all ongoing infections at the end of follow-up were classified into the unknown/ongoing group. All infections with lingering symptoms, such as Long COVID or long-term loss of smell/taste were classified into the lingering category.
7. N=218: Includes only participants who experienced a SARS-CoV-2 infection and an additional 5 participants were removed due to unknown dyspnoea severity/duration.
8. Symptoms were reported on a scale of 0-5, 0 for not present and 1-5 present with increasing severity. Fever was reported as present or not and was defined as temperatures above 38°C.
9. Respiratory symptoms exclude dyspnoea. Most participants reported more than one type of respiratory symptom during an infection episode, therefore we took the highest severity reported for any respiratory symptom. Accordingly, the duration was calculated from the day on which the first respiratory symptom was reported until the day the last respiratory symptom disappeared.
10. N=205: Includes only participants who experienced a SARS-CoV-2 infection and an additional 18 were removed due to unknown respiratory symptom severity.
11. N=200: Includes only participants who experienced a SARS-CoV-2 infection and an additional 23 were removed due to unknown or ongoing respiratory symptom duration.
12. N=217: Includes only participants who experienced a SARS-CoV-2 infection and an additional 6 were removed due to unknown presence of fever or unknown fever duration.
13. N=205: Includes only participants who experienced a SARS-CoV-2 infection and an additional 18 were removed due to unknown non-respiratory symptom severity.
14. Non-respiratory symptoms exclude fever. Most participants reported more than one type of non-respiratory symptom during an infection episode, and we therefore took the highest severity reported for any non-respiratory symptom. Accordingly, the duration was calculated from the day on which the first non-respiratory symptom appeared until the day the last non-respiratory symptom disappeared.

**Figure S7: Sensitivity analyses: adding covariates to the final multivariable regression models**

**A. Multivariable model with M12 anti-S1 log_10_ concentrations as outcome^1^**

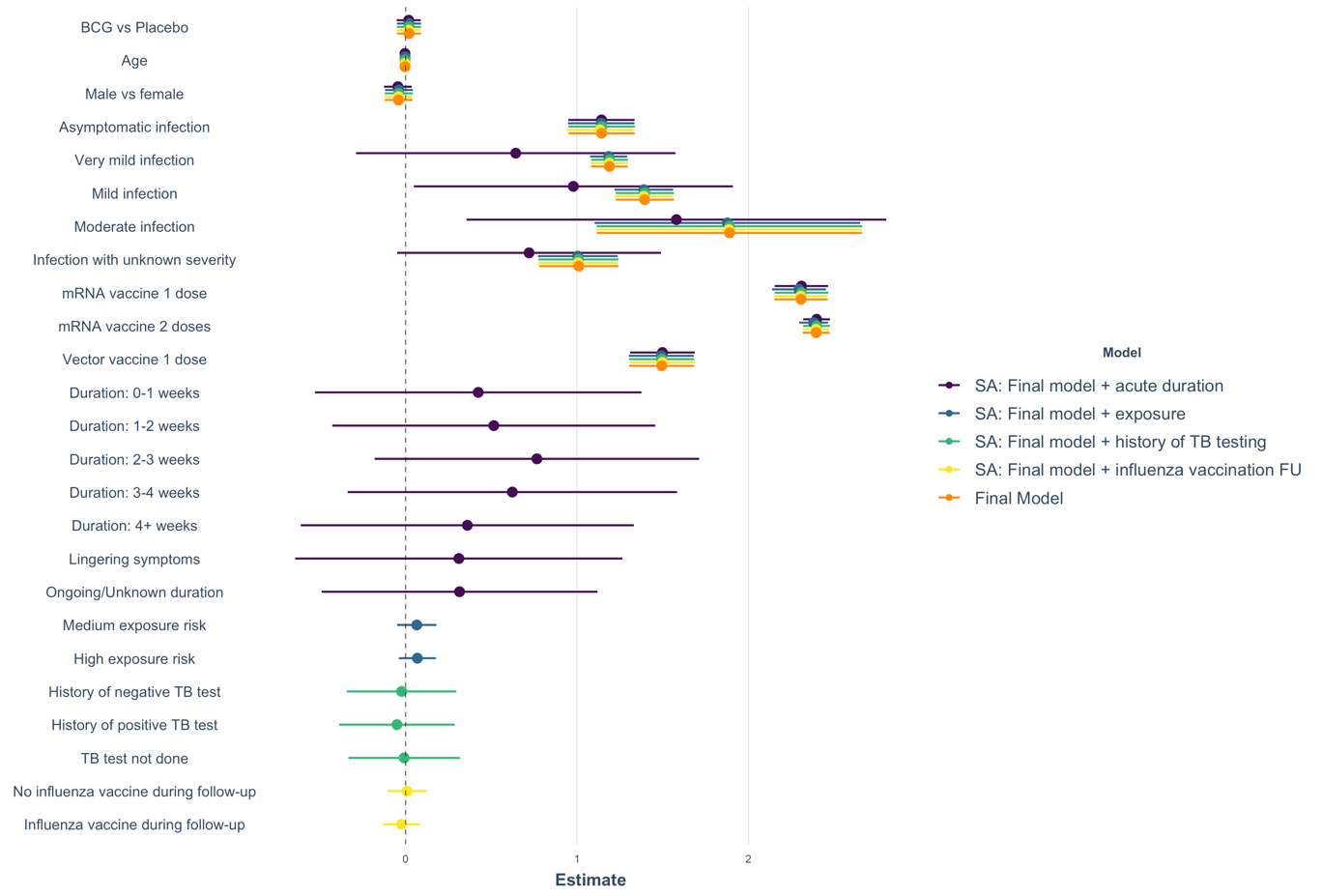

**B. Multivariable model with M12 anti-N log_10_ concentrations as outcome^1^**

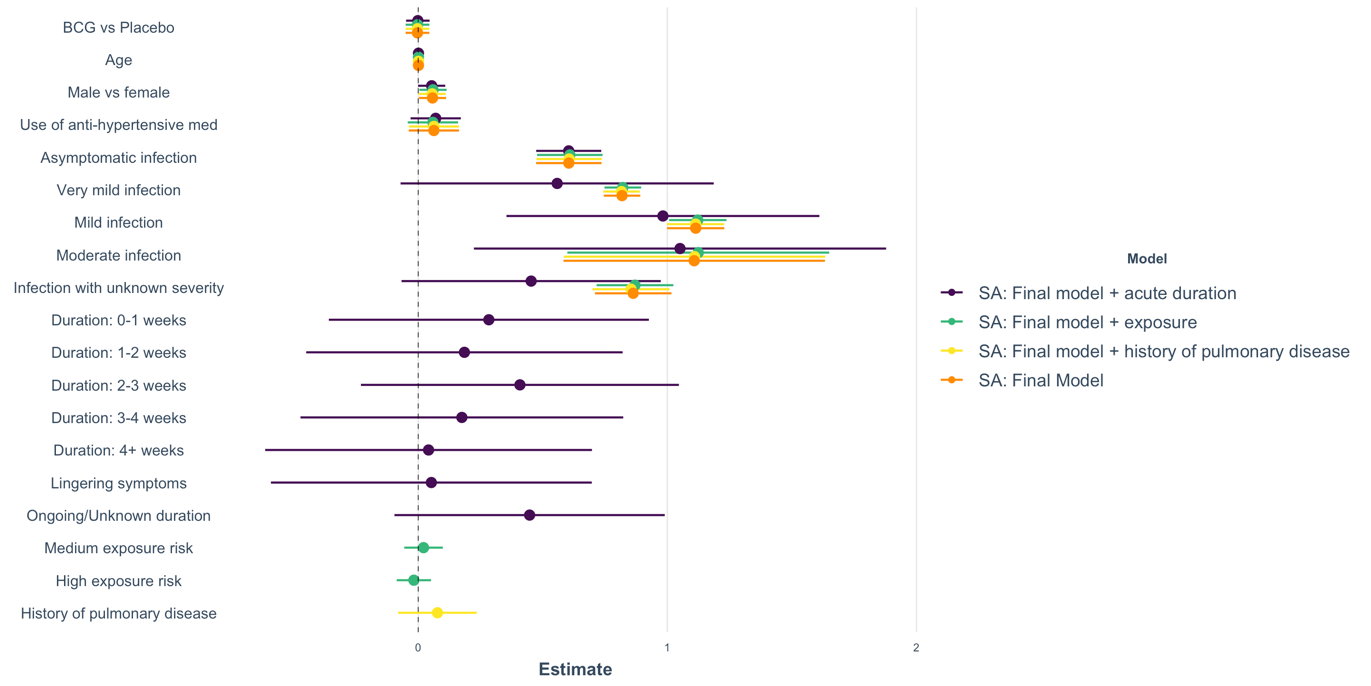

Abbreviations: BCG=Bacillus Calmette-Guérin; FU=follow up; med=medication; SA=sensitivity analysis; TB=tuberculosis.

1.	The reference is an individual in the placebo group, female, who never experienced aSARS-CoV-2 infection and never received a single dose of a COVID-19 vaccine. Age is continuous (per year). The reference for workplace exposure risk is low risk. The reference for past TB test results is only having had negative TB test results.

**Table S4: Sensitivity analyses: linear regression models after applying varying seroconversion windows**

**A: Multivariable model^1^ with M12 anti-S1 log_10_ concentrations as outcome**

| **Covariate** | **Seroconversion window = 14 days^3,4^** | | **Seroconversion window = 7 days^3,5^** | | **Seroconversion window = 0 days^3,6^** | |
| --- | --- | --- | --- | --- | --- | --- |
|  | **Estimate (95% CI)** | **p-value** | **Estimate (95% CI)** | **p-value** | **Estimate (95% CI)** | **p-value** |
| Model intercept | 0.50 (0.36, 0.64) | **<0.001** | 0.47 (0.33, 0.61) | **<0.001** | 0.44 (0.28, 0.60) | **<0.001** |
| BCG vs placebo^2^ | 0.02 (-0.05, 0.09) | 0.585 | 0.06 (-0.01, 0.13) | 0.118 | 0.02 (-0.06, 0.10) | 0.627 |
| Age (per year) | -0.004 (-0.01, -0.007) | **0.008** | -0.004 (-0.01, 0.001) | **0.009** | -0.004 (-0.01, -0.001) | **0.034** |
| Male sex | -0.04 (-0.12, 0.04) | 0.311 | -0.0 (-0.11, 0.06) | 0.598 | 0.01 (-0.08, 0.11) | 0.773 |
| **All the below compared to participants who never had a SARS-CoV-2 infection or COVID-19 vaccination during follow-up:** | | | | | | |
| Asymptomatic | 1.14 (0.95, 1.34) | **<0.001** | 1.07 (0.87, 1.27) | **<0.001** | 1.11 (0.88, 1.33) | **<0.001** |
| Very mild | 1.19 (1.08, 1.30) | **<0.001** | 1.25 (1.14, 1.37) | **<0.001** | 1.25 (1.13, 1.37) | **<0.001** |
| Mild | 1.39 (1.22, 1.56) | **<0.001** | 1.48 (1.31, 1.66) | **<0.001** | 1.66 (1.47, 1.85) | **<0.001** |
| Moderate | 1.89 (1.12, 2.66) | **<0.001** | 2.16 (1.47, 2.84) | **<0.001** | 2.71 (1.94, 3.48) | **<0.001** |
| Unknown severity | 1.01 (0.78, 1.24) | **<0.001** | 1.13 (0.88, 1.37) | **<0.001** | 1.12 (0.85, 1.40) | **<0.001** |
| mRNA 1 dose | 2.31 (2.15, 2.46) | **<0.001** | 1.86 (1.71, 2.00) | **<0.001** | 1.04 (0.91, 1.18) | **<0.001** |
| mRNA 2 doses | 2.40 (2.32, 2.47) | **<0.001** | 2.40 (2.31, 2.48) | **<0.001** | 2.41 (2.32, 2.50) | **<0.001** |
| Vector 1 dose | 1.49 (1.31, 1.68) | **<0.001** | 1.28 (1.11, 1.45) | **<0.001** | 0.87 (0.71, 1.03) | **<0.001** |
| **Model fit** | **R^2^=0.84** | | **R^2^=0.81** | | **R^2^=0.76** | |

**B: Multivariable model^1^ with M12 anti-N log_10_ concentrations as outcome**

| **Covariate** | **Seroconversion window = 14 days^3,4^** | | **Seroconversion window = 7 days^3,5^** | | **Seroconversion window = 0 days^3,6^** | |
| --- | --- | --- | --- | --- | --- | --- |
|  | **Estimate (95% CI)** | **p-value** | **Estimate (95% CI)** | **p-value** | **Estimate (95% CI)** | **p-value** |
| Model intercept | 0.48 (0.39, 0.57) | **<0.001** | 0.47 (0.38, 0.56) | **<0.001** | 0.47 (0.38, 0.55) | **<0.001** |
| BCG vs placebo^2^ | -0.003 (-0.05, 0.04) | 0.901 | -0.004 (-0.05, 0.05) | 0.860 | -0.004 (-0.05, 0.04) | 0.876 |
| Age (per year) | 0.001 (-0.007, 0.003) | 0.312 | 0.001 (-0.001, 0.003) | 0.224 | 0.001 (-0.001, 0.003) | 0.267 |
| Male sex | 0.06 (0.005, 0.11) | **0.040** | 0.06 (0.01, 0.12) | **0.018** | 0.06 (0.01, 0.12) | **0.019** |
| Hypertension med | 0.06 (-0.04, 0.16) | 0.222 | 0.05 (-0.04, 0.15) | 0.284 | 0.05 (-0.04, 0.15) | 0.287 |
| **All the below compared to participants who never had a SARS-CoV-2 infection or COVID-19 vaccination during follow-up:** | | | | | | |
| Asymptomatic | 0.60 (0.47, 0.74) | **<0.001** | 0.63 (0.50, 0.76) | **<0.001** | 0.62 (0.49, 0.74) | **<0.001** |
| Very mild | 0.82 (0.74, 0.89) | **<0.001** | 0.82 (0.75, 0.89) | **<0.001** | 0.81 (0.74, 0.87) | **<0.001** |
| Mild | 1.11 (1.00, 1.23) | **<0.001** | 1.07 (0.96, 1.18) | **<0.001** | 1.07 (0.96, 1.18) | **<0.001** |
| Moderate | 1.11 (0.58, 1.63) | **<0.001** | 1.10 (0.67, 1.52) | **<0.001** | 1.11 (0.68, 1.54) | **<0.001** |
| Unknown severity | 0.86 (0.71, 1.02) | **<0.001** | 0.88 (0.73, 1.03) | **<0.001** | 0.88 (0.73, 1.03) | **<0.001** |
| **Model fit** | **R^2^=0.49** | | **R^2^=0.49** | | **R^2^=0.48** | |

Abbreviations: CI=Confidence interval; M=month; med=medication.

1. The reference is an individual in the placebo group, female, who never experienced a SARS-CoV-2 infection and never received a single dose of a COVID-19 vaccine.
2. BCG vs placebo vaccination at baseline was forced into the model to take the randomisation into account.
3. The VIF values for this model are all less than 5, which means that there is no evidence of multicollinearity.
4. Window period is 14 days. The model includes 970 participants and 223 infections. Four participants were in the seroconversion window for an infection and 125 for the first dose of a vaccine.
5. Window period is 7 days. The model includes 1,024 participants and 236 infections. Four participants were in the seroconversion window for an infection and 71 for the first dose of a vaccine.
6. Window period is 0 days. The model includes 1,099 participants and 249 infections.
